# COVID-19 collateral: Indirect acute effects of the pandemic on physical and mental health in the UK

**DOI:** 10.1101/2020.10.29.20222174

**Authors:** Kathryn E Mansfield, Rohini Mathur, John Tazare, Alasdair D Henderson, Amy Mulick, Helena Carreira, Anthony A Matthews, Patrick Bidulka, Alicia Gayle, Harriet Forbes, Sarah Cook, Angel YS Wong, Helen Strongman, Kevin Wing, Charlotte Warren-Gash, Sharon L Cadogan, Liam Smeeth, Joseph F Hayes, Jennifer K Quint, Martin McKee, Sinéad M Langan

## Abstract

**Background:** Concerns have been raised that the response to the UK COVID-19 pandemic may have worsened physical and mental health, and reduced use of health services. However, the scale of the problem is unquantified, impeding development of effective mitigations. We asked what has happened to general practice contacts for acute physical and mental health outcomes during the pandemic?

**Methods:** Using electronic health records from the Clinical Research Practice Datalink (CPRD) Aurum (2017-2020), we calculated weekly primary care contacts for selected acute physical and mental health conditions (including: anxiety, depression, acute alcohol-related events, asthma and chronic obstructive pulmonary disease [COPD] exacerbations, cardiovascular and diabetic emergencies). We used interrupted time series (ITS) analysis to formally quantify changes in conditions after the introduction of population-wide restrictions (‘lockdown’) compared to the period prior to their introduction in March 2020.

**Findings:** The overall population included 9,863,903 individuals on 1^st^ January 2017. Primary care contacts for all conditions dropped dramatically after introduction of population-wide restrictions. By July 2020, except for unstable angina and acute alcohol-related events, contacts for all conditions had not recovered to pre-lockdown levels. The largest reductions were for contacts for: diabetic emergencies (OR: 0.35, 95% CI: 0.25-0.50), depression (OR: 0.53, 95% CI: 0.52-0.53), and self-harm (OR: 0.56, 95% CI: 0.54-0.58).

**Interpretation:** There were substantial reductions in primary care contacts for acute physical and mental conditions with restrictions, with limited recovery by July 2020. It is likely that much of the deficit in care represents unmet need, with implications for subsequent morbidity and premature mortality. The conditions we studied are sufficiently severe that any unmet need will have substantial ramifications for the people experiencing the conditions and healthcare provision. Maintaining access must be a key priority in future public health planning (including further restrictions).

**Funding:** Wellcome Trust Senior Fellowship (SML), Health Data Research UK.

**RESULTS IN CONTEXT:** *Evidence before this study:* A small study in 47 GP practices in a largely deprived, urban area of the UK (Salford) reported that primary care consultations for four broad diagnostic groups (circulatory disease, common mental health problems, type 2 diabetes mellitus and malignant cancer) declined by 16-50% between March and May 2020, compared to what was expected based on data from January 2010 to March 2020. We searched Medline for other relevant evidence of the indirect effect of the COVID-19 pandemic on physical and mental health from inception to September 25^th^ 2020, for articles published in English, with titles including the search terms (“covid*” or “coronavirus” or “sars-cov-2”), and title or abstracts including the search terms (“indirect impact” or “missed diagnos*” or “missing diagnos*” or “delayed diagnos*” or ((“present*” or “consult*” or “engag*” or “access*”) AND (“reduction” or “decrease” or “decline”)). We found no further studies investigating the change in primary care contacts for specific physical- and mental-health conditions indirectly resulting from the COVID-19 pandemic or its control measures. There has been a reduction in hospital admissions and presentations to accident and emergency departments in the UK, particularly for myocardial infarctions and cerebrovascular accidents. However, there is no published evidence specifically investigating the changes in primary care contacts for severe acute physical and mental health conditions.

*Added value of this study:* To our knowledge this is the first study to explore changes in healthcare contacts for acute physical and mental health conditions in a large population representative of the UK. We used electronic primary care health records of nearly 10 million individuals across the UK to investigate the indirect impact of COVID-19 on primary care contacts for mental health, acute alcohol-related events, asthma/chronic obstructive pulmonary disease (COPD) exacerbations, and cardiovascular and diabetic emergencies up to July 2020. For all conditions studied, we found primary care contacts dropped dramatically following the introduction of population-wide restriction measures in March 2020. By July 2020, with the exception of unstable angina and acute alcohol-related events, primary care contacts for all conditions studied had not recovered to pre-lockdown levels. In the general population, estimates of the absolute reduction in the number of primary care contacts up to July 2020, compared to what we would expect from previous years varied from fewer than 10 contacts per million for some cardiovascular outcomes, to 12,800 per million for depression and 6,600 for anxiety. In people with COPD, we estimated there were 43,900 per million fewer contacts for COPD exacerbations up to July 2020 than what we would expect from previous years.

*Implicatins of all the available evidence:* While our results may represent some genuine reduction in disease frequency (e.g. the restriction measures may have improved diabetic glycaemic control due to more regular daily routines at home), it is more likely the reduced primary care conatcts we saw represent a substantial burden of unmet need (particularly for mental health conditions) that may be reflected in subsequent increased mortality and morbidity. Health service providers should take steps to prepare for increased demand in the coming months and years due to the short and longterm ramifications of reduced access to care for severe acute physical and mental health conditions. Maintaining access to primary care is key to future public health planning in relation to the pandemic.

## INTRODUCTION

By October 2020, novel coronavirus disease 2019 (COVID-19) has been diagnosed in more than 40 million individuals, with over one million deaths reported worldwide.^1^ Much research and public health attention has, understandably, focused on preventing infections and reducing mortality. However, there are concerning reports of decreased health service use.^2–5^ Inevitably, there will be impacts on *non*-COVID-19-related healthcare provision; healthcare resources have been reallocated to the COVID-19 response and care delivery has been modified due to mitigation measures including social distancing.^6–11^ Additionally, individuals may have delayed seeking care (due to fear of infection, or to avoid burdening health services). Psychological health will have been affected by pandemic-related fears, employment and financial concerns, and control measures (including social distancing, closures of social spaces and isolation),^12,13^ and lockdown measures are likely to have reduced access to mental health care (face-to-face visits and talking therapies). Understanding the indirect effects of the pandemic and its control measures is essential for public health planning, particularly when/if the COVID-19 pandemic is under control (or if further restrictions are needed), and for informing control measures for future pandemics.

Reports indicate that accident and emergency department attendance and hospital admissions for non-COVID-related acute concerns in the UK have declined since March 2020.^2–4^ However, it is not yet clear what has happened in primary care across the UK where clinical work has changed rapidly to include more remote consultations,^14–17^ although a regional report indicates reduced primary care consultations.^18^

We asked how primary care contacts (including face-to-face or remote consultations, and recording of diagnoses from hospital discharge summaries) have changed for selected indirect acute physical and mental health effects of the COVID-19 pandemic, to inform decisions on policy responses and resource allocation. Although a wide range of diagnoses could be indirectly affected by the COVID-19 pandemic, we focused on specific acute conditions that could plausibly be affected including: mental health conditions, acute alcohol-related events, cardiovascular and diabetic emergencies, and asthma and chronic obstructive pulmonary disease (COPD) exacerbations. We specifically selected diabetic and cardiovascular emergencies (including myocardial infarction, unstable angina), and asthma/COPD exacerbations, as affected individuals are likely to be considered vulnerable (and asked to ‘shield’, i.e. advice for the vulnerable to avoid unnecessary contacts, to avoid infection),^19^ creating a barrier to accessing healthcare resources.

## METHODS

We used routinely collected primary care data from electronic health records from general practices contributing to Clinical Research Practice Datalink (CPRD) Aurum database (August 2020 build) in the three years prior to the COVID-19 pandemic and four months after introducing population-wide restrictions (i.e. ‘lockdown’) on 23^rd^ March 2020 (1^st^ January 2017-12^th^ July 2020).^20^ Code lists for defining all outcomes and stratifying variables, and analytic code are available (https://github.com/johntaz/COVID-Collateral).

### Data source

CPRD Aurum includes de-identified routinely collected primary care health record data from participating general practices covering 13% of the UK population, and is broadly representative of the English population with respect to age, sex, ethnicity, and geographic region.^20^

### Study population

Our overall study population included individuals with at least one year of registration with practices contributing to CPRD Aurum (January 2017-July 2020). Included populations (i.e. denominators) varied depending on the condition being investigated (**Table 1, Figure S1**). For example, for diabetic emergencies the study population (denominator) only included individuals (aged ≥11 years) with an existing diabetes mellitus diagnosis, while for depression, the study included all individuals from the overall study population aged 5 years and older.

**Table 1.** Description of denominator populations and condition definitions.

| Condition | Denominator population (condition-specific denominator populations) | Condition definition |
| --- | --- | --- |
| **Diabetes** |  |  |
| Diabetic emergencies | All individuals (aged ≥11 years) with prevalent diagnoses of diabetes mellitus at the start of each week of follow-up. Individuals contributed to the study population from the latest of the start of follow-up in the overall population and the date of their first record indicating a diagnosis of diabetes. | Any record of diabetes-related hyperglycaemia, hypoglycaemia, ketoacidosis, or diabetic coma. Multiple records occurring within **seven days** of each other were considered as representing the same event. |
| **Mental health** |  |  |
| Anxiety | All individuals (aged ≥11 years) from the overall study population. | Any record of symptoms or diagnoses of: social phobia, agoraphobia, panic, generalized anxiety disorder, and mixed anxiety and depression. Multiple records occurring within **seven days** of each other were considered as representing the same event. |
| Depression | All children (aged 5-17 years) and adults (aged ≥18) from the overall study population. | Any record of major depressive disorder, dysthymia, mixed anxiety and depression, and adjustment disorders with depressed mood. We also included codes for depressive symptoms. Multiple records occurring within **seven days** of each other were considered as representing the same event. |
| Self-harm | All children (aged 5-17 years) and adults (aged ≥18 years) from the overall study population. | Self-harm defined as records that indicated explicit or undertermined intention to self-harm, non-suicidal or suicidal self-harm (including overdoses with drugs commonly implicated in suicide, e.g. paracetamol). Multiple records occurring within **seven days** of each other were considered as representing the same event. |
| Serious mental illness | All children (aged 5-17 years) and adults (aged ≥18 years) from the overall study population. | Severe mental illness included diagnoses of schizophrenia and other psychotic disorders, and bipolar disorders. Multiple records occurring within **seven days** of each other were considered as representing the same event. |
| Eating disorders | All children (aged 5-17 years) and adults (aged ≥18 years) from the overall study population. | Eating disorders included anorexia nervosa, bulimia nervosa, and other specified feeding and eating disorders. Multiple records occurring within **seven days** of each other were considered as representing the same event. |
| Obsessive compulsive disorder | All children (aged 5-17 years) and adults (aged ≥18 years) from the overall study population. | Obsessive compulsive disorder was defined by codes for body dysmorphic disorders, hypochondriasis, hoarding disorder, and body focused repetitive behaviour disorders. Multiple records occurring within **seven days** of each other were considered as representing the same event. |
| **Respiratory** |  |  |
| Asthma exacerbations | All individuals (aged $\geq 11$ years) with a current asthma diagnosis (i.e. asthma code in the last two or three years if aged $< 18$ years or $18+$ years, respectively). Individuals joined the study population from the start of follow-up in the overall population if there was a current asthma diagnosis (i.e. within last 2-3 years) at this time or from the date of their first record indicating an asthma diagnosis within overall follow-up. Participants remained in the study until there was no current asthma diagnosis or the end of overall follow-up. They were able to re-enter the study if there was a later diagnostic code for asthma before the end of overall follow-up. Following an existing definition, individuals 40 years and over with asthma were considered as likely to have COPD (and therefore not included in the asthma study population [denominator]) if they had a subsequent COPD diagnosis recorded within the two years following the current asthma record.<sup>74</sup> | Asthma exacerbations were defined as records for morbidity codes for asthma exacerbations and status asthmaticus, and a primary care prescription for an oral corticosteroid.<sup>21</sup> Multiple records occurring within **14 days** of each other were considered as representing the same event. |
| COPD exacerbations | Adults (aged $\geq 41$ years) with an established diagnosis of COPD and evidence of a smoking history.<sup>75</sup> Individuals joined the study population from the latest of the start of follow-up in the overall population and the date of their first record indicating diagnosis of COPD. | Exacerbations of COPD were defined using morbidity codes in individuals with existing COPD for COPD exacerbations, lower respiratory tract infections, breathlessness or sputum production, and a new prescription for an oral corticosteroid or antibiotic.<sup>22</sup> Multiple records occurring within **14 days** of each other were considered as representing the same event. |
| **Cardiovascular** |  |  |
| Myocardial infarction | All adults (aged $> 31$ years) | Any record for myocardial infarction allowing for a **1-year** window between successive records. Multiple records occurring within one year of each other were considered as representing the same event. |
| Unstable angina | All adults (aged $\geq 31$ years) | Any record for unstable angina, allowing for a **6-month** window between successive records. Multiple records occurring within six months of each other were considered as representing the same event. |
| Transient ischaemic attacks | All adults (aged $\geq 31$ years) | Any record for transient ischaemic, allowing for a **6-month** window between successive records. Multiple records occurring within six months of each other were considered as representing the same events |
| Cerebrovascular accident | All adults (aged $\geq 31$ years) | Any record for cerebrovascular accidents, allowing for a **1-year** window between successive records. Multiple records occurring within one year of each other were considered as representing the same event. |
| Cardiac failure | All adults (aged ≥31 years) | Given the complexity with capturing acute events for a chronic condition, we only counted an individual's **first ever diagnosis** with cardiac failure. |
| Venous thromboembolism (pulmonary embolism and deep venous thrombosis) | All adults (aged ≥31 years) | Any record for venous thromboembolism, allowing for a **1-year** window between successive records. Multiple records occurring within one year of each other were considered as representing the same event. |
| **Alcohol** |  |  |
| Acute alcohol-related event | All adults (aged ≥18 years) | Any record for acute physical or psychological alcohol-related event, including acute alcoholic pancreatitis. Multiple records occurring within **14 days** of each other were considered as representing the same event. |

- 74 Bloom CI, Nissen F, Douglas IJ, Smeeth L, Cullinan P, Quint JK. Exacerbation risk and characterisation of the UK's asthma population from infants to old age. *Thorax* 2018; 73: 313–20. - 21 Bloom CI, Palmer T, Feary J, Quint JK, Cullinan P. Exacerbation patterns in adults with Asthma in England A population-based study. *Am J Respir Crit Care Med* 2019; 199: 446–53. - 75 Quint JK, Müllerova H, DiSantostefano RL, et al. Validation of chronic obstructive pulmonary disease recording in the Clinical Practice Research Datalink (CPRD-GOLD). *BMJ Open* 2014; 4: 1–8. - 22 Rothnie KJ, Müllerová H, Hurst JR, et al. Validation of the recording of acute exacerbations of COPD in UK primary care electronic healthcare records. *PLoS One* 2016; 11: 1–14.

We followed all individuals from the latest of: study start (1^st^ January 2017), one year from GP registration or, for diabetes and respiratory conditions, from meeting our definitions for having diabetes or respiratory disease, as appropriate (**Table 1**). Follow-up ended for all study populations at the earliest of: end of registration with GP, death, practice stopped contributing to CPRD, or end of the study period.

### Exposures, outcomes and stratifying variables

Our exposure was the introduction of population-wide COVID-19 restrictions (i.e. ‘lockdown’ on 23^rd^ March 2020). As outcomes, we considered the number of weekly primary care contacts for the following conditions (separately): mental health (i.e. depression, anxiety, fatal and non-fatal self-harm, severe mental illness, and eating and obsessive-compulsive disorders), acute alcohol-related events, diabetic emergencies (e.g. ketoacidosis), asthma and COPD exacerbations, and acute cardiovascular (CVD) events (i.e. unstable angina, myocardial infarction, transient ischaemic attack, cerebrovascular accident, cardiac failure and venous thromboembolisms). We used the term ‘contact’ broadly to represent remote and face-to-face consultations, diagnoses from hospital discharge letters, and secondary care referrals. We identified conditions using primary care records for diagnoses, symptoms and/or prescribing (**Table 1**). All outcomes, except asthma/COPD exacerbations, were captured based on presence/absence of specific morbidity codes. Asthma/COPD exacerbations were based on validated algorithms requiring a combination of specific morbidity codes and prescriptions for corticosteroids or, for COPD additionally antibiotics.^21,22^ For some conditions we defined an exclusion period during which we regarded further coding for the same outcome as representing the same acute event (e.g. for diabetic emergencies we regarded multiple records within seven days of each other as representing the same event). We used different condition-specific time periods to define outcome events to account for differences in natural history of study outcomes (**Table 1**).

We stratified on the following pre-specified variables: age (in 10-year bands), sex, geographic region, and ethnicity.

### Statistical analysis

We described all denominator study populations in the first week of January each year (2017-2020). We plotted the percentage of our study populations with contacts for particular conditions in given weeks in 2020 and historical averages for that week (2017-2019). We repeated analyses stratified by age, sex, region, and ethnicity.

To quantify changes in consultation behaviour following introducing restrictions we used an interrupted time series (ITS) analysis separating our time series into two periods: 1) Pre-lockdown: 1^st^ January 2017 to March 1^st^ 2020 for all outcomes except self-harm (which excluded data from 2017 and 2018, **Text S1**); and 2) With-restrictions: from 29^th^ March 2020 to the study end (12^th^ July 2020).

Although restrictions were announced on 23^rd^ March,^23^ activity levels (measured by mobile phone applications and public transport journeys) had declined before the announcement.^24–26^ To account for anticipatory behaviour, we conservatively defined the start of restrictions as the first week of March, and removed all data in March up to, and including, the week restrictions were announced from the ITS analysis.

For our ITS analysis, we used binomial generalised linear models with number of weekly contacts weighted by dynamic population sizes (updated weekly).^27^ We included a linear effect of time to capture long-term behaviour trends, a binary pre-lockdown/with-restrictions variable to measure the direct ‘step’ change in behaviour, and an interaction between the two to allow for a recovery ‘slope’ change in behaviour. We accounted for seasonal effects by including calendar month as a categorical variable, and autocorrelation by including first order lagged residuals. Standard errors were scaled to account for overdispersion.^27^

To estimate the reduction in contacts as restrictions were introduced (the ‘step’ change) we reported odds ratios (ORs) for the relative difference in contacts at the start of the with-restrictions period compared to the end of the pre-lockdown period. To estimate the recovery of contacts over time (the ‘slope’) we used the coefficients from the ITS model to estimate the weekly log odds of contact during the with-restrictions period (further details: **Text S2**).

To estimate absolute effects of restrictions on the number of contacts, we repeated our analysis using Poisson regression to generate linear predictions of the estimated log contact count and the estimated log count if the ‘restrictions’ term was set to zero (i.e. there had been no restrictions). To quantify absolute changes in behaviour over time we compared the point estimate of the estimated number of contacts with and without restrictions at two time points: 1 month (26^th^ April) and 3 months after introduction of restrictions (28^th^ June).

We used Stata version 16^28^ and R version 4.0.2^29^ for our analyses.

### Sensitivity analyses

Our definitions for pre-lockdown and with-restrictions periods may have influenced our estimates, so we repeated the ITS analysis with the same pre-lockdown period but with variable data-exclusion periods (5 and 7 weeks, versus 3 weeks in the main analysis) and repeated analyses with the pre-lockdown period ending on 15^th^ March, the week before restriction introduction,^23^ excluding data for 0, 3, 5 and 7 weeks as sensitivity analyses. Additionally, given the small number of diabetic emergency contacts we varied our definition using less specific codes in a *post-hoc* sensitivity analysis (**Text S3**).

## RESULTS

The overall denominator population included 9,863,903 individuals on 1^st^ January 2017 and numbers remained relatively stable throughout the study (**Table 2**). Characteristics of condition-specific study populations are in the Appendix (**Tables S1-S5**).

**Table 2.**
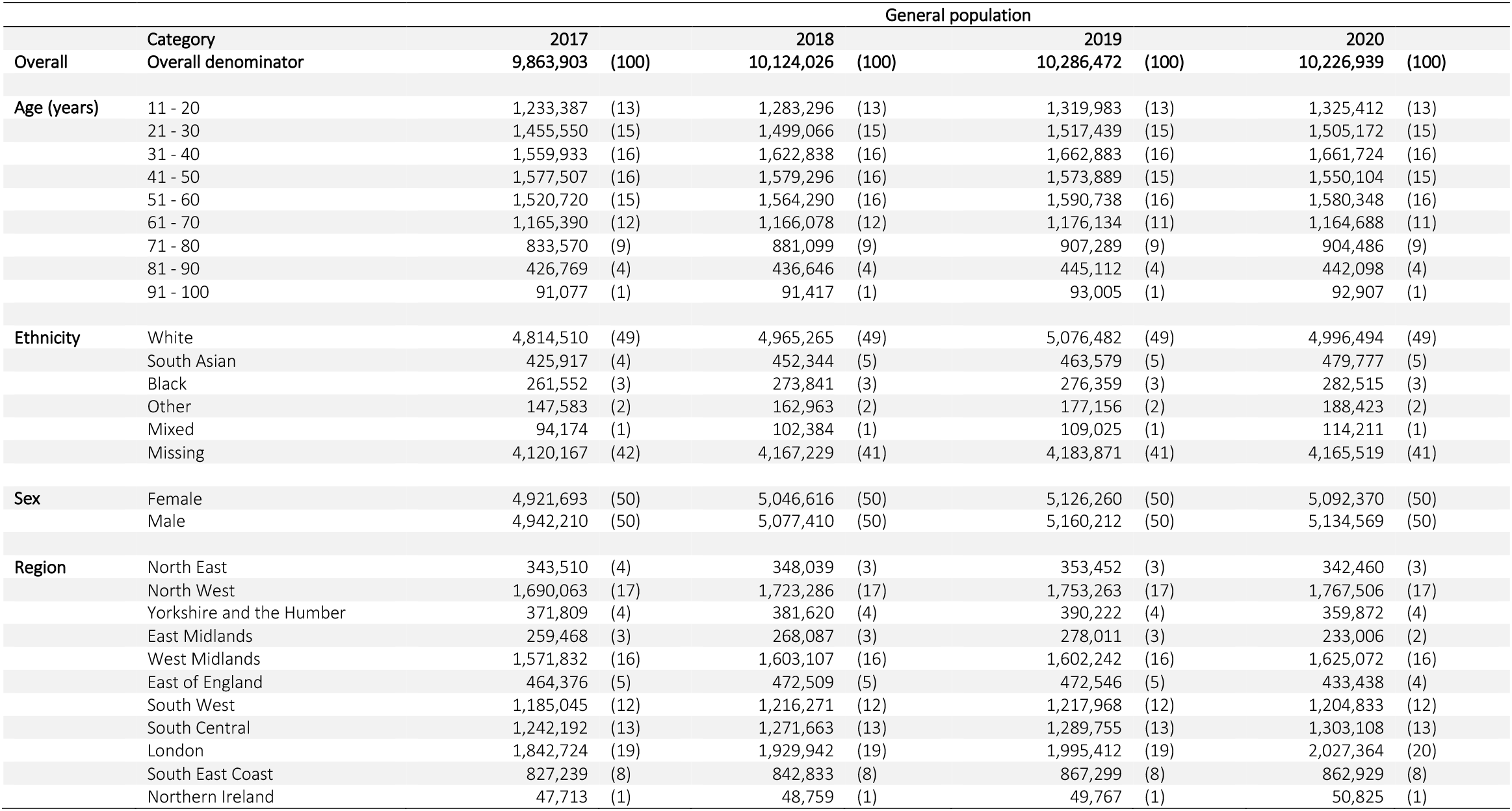
General population denominator population defined in first week of January 2017-2020. All figures are n (%)

**Figure 1** shows the percentage of a given study populations with primary care contacts for each condition in 2020 (red line) and a 3-year historical average for the corresponding week (black line). Across the majority of conditions, we observed rapid and sustained decreases in GP contacts between March and July 2020 compared to pre-lockdown periods. Despite gradual increases in the percentage of contacts following restrictions, levels remained below the three-year average for all conditions except acute alcohol-related events, which were higher than the historical average in 2020, and unstable angina. During March 2020 we observed pronounced increases in contacts related to asthma exacerbations. Patterns were broadly consistent when stratified by age (**Figure 2**), sex, region, and ethnicity (**Figures S2-S4**).

**Figure 1.**
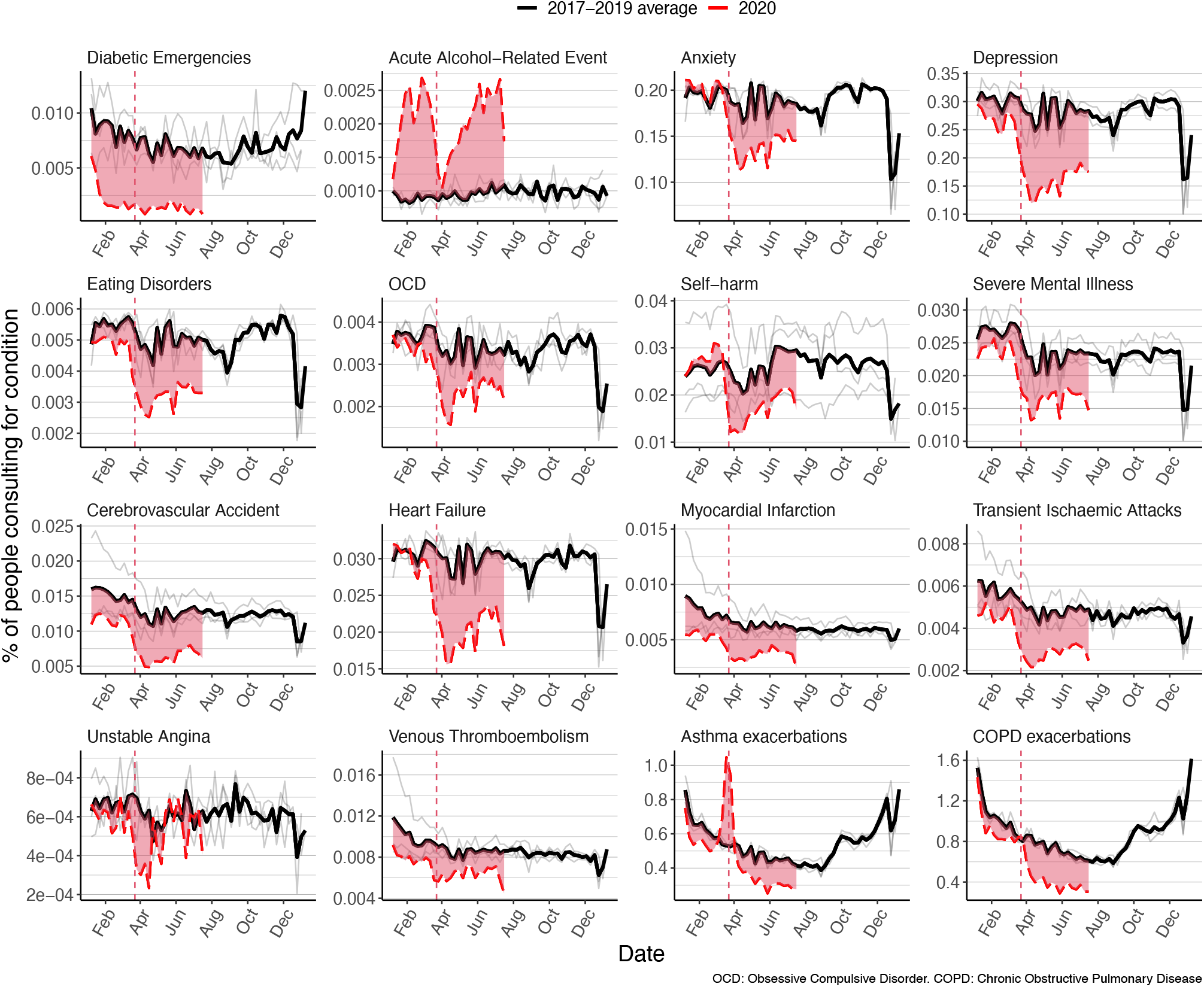
Percentage of study populations with contacts for all conditions in 2017-2019 and 2020. Percentage of eligible population with contacts for each health condition studied in 2020 compared to historical average for that week. Black line, weekly historical average percentage of eligible population consulting (2017-2019, grey lines show the data for 2017, 2018, and 2019). Red dashed line, weekly percentage of eligible percentage consulting in 2020. Red region shows difference with historical average. Red dotted line, introduction of restrictions in UK on March 23^rd^

**Figure 2.**
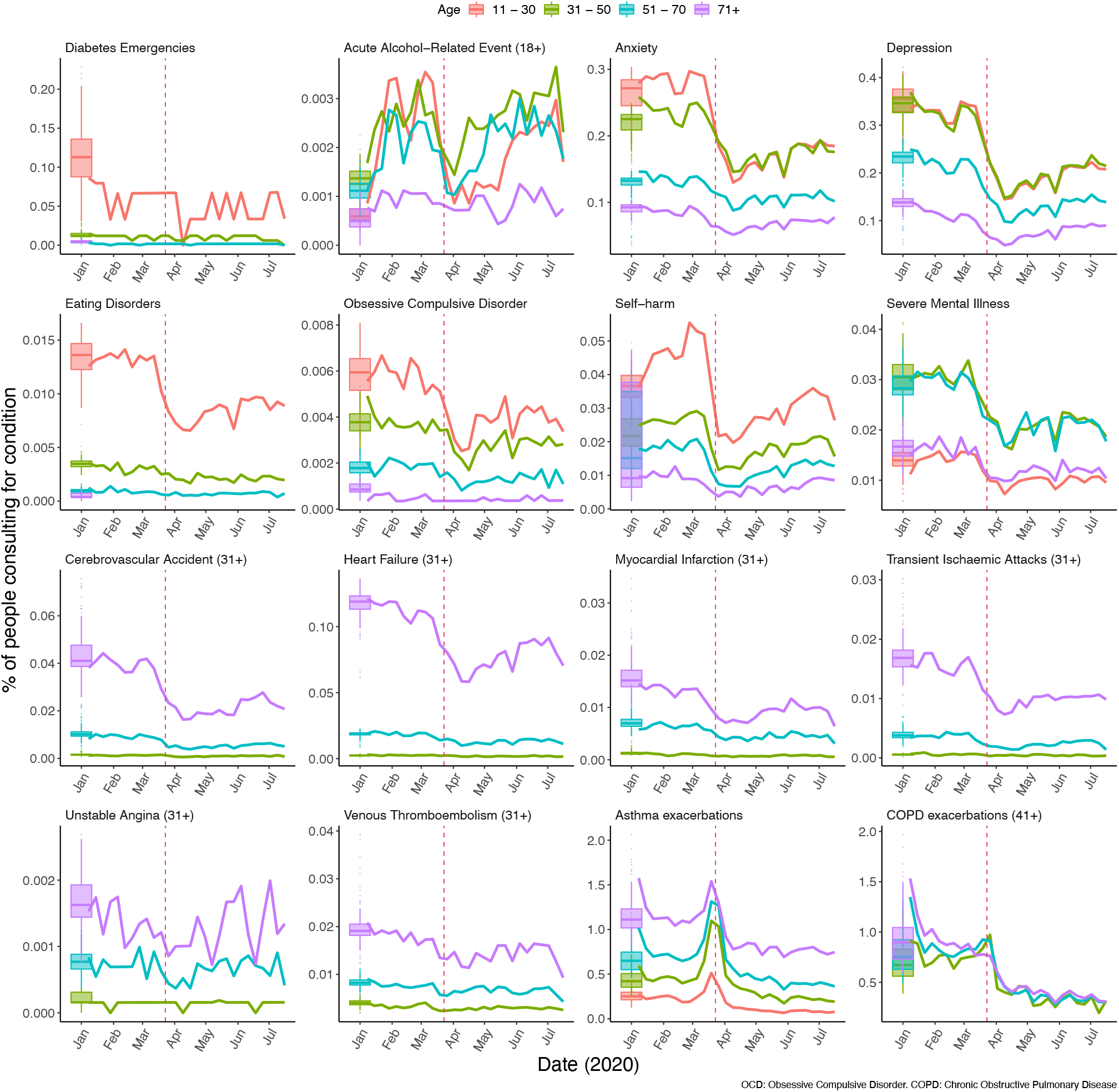
Percentage of the study populations with contacts by age groupd. Percentage of the study populations with GP contacts for study conditions over 2020, by age group. Boxplots, historical average percentage of study population with GP contacts for the condition of interest. Coloured lines, weekly percentage of eligible population with primary care contacts for the condition of interest in 2020. Red dotted line, introduction of restrictions in UK on March 23^rd^. Note that cell counts with fewer than five contacts in one week in 2020 have been suppressed.

There was evidence that contacts for all studied conditions, except acute alcohol-related events, were lower after restrictions were announced compared to pre-restriction levels (**Figure 3A**). The largest relative reductions in contact behaviour following restriction introduction were observed for diabetic emergencies (OR: 0.35, 95% CI: 0.25-0.50), depression (OR: 0.53, 95% CI: 0.52-0.53), and self-harm (OR: 0.56, 95% CI: 0.54-0.58) (**Figure 3B, Table S6**). From March 28^th^ 2020, we saw evidence of increasing contacts for most conditions over time. Acute alcohol-related events and unstable angina contacts appeared to recover faster (3-5% increase in odds of contact per week) (**Figure 3C, Table S7**) than, for example, mental health contacts, where odds of contact increased by 1-2% per week, despite a 20-40% drop following restrictions (**Figure 3B, Table S6**). Sensitivity analyses varying the choice of restriction period provided broadly consistent results over a wide range of scenarios with a notable exception of acute alcohol-related events (**Tables S6-S7, Figures S8-S14**).

**Figure 3.**
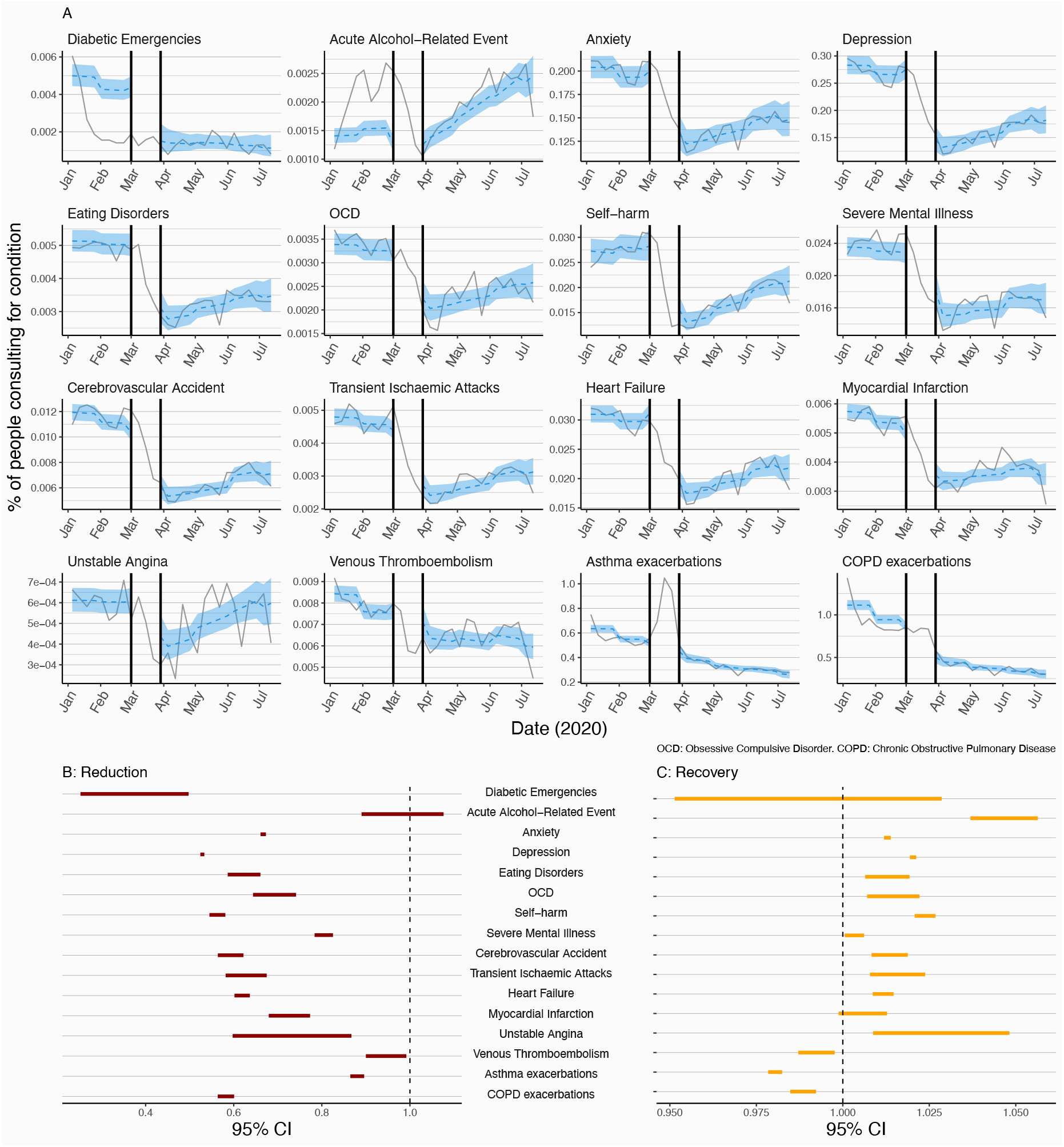
Interrupted time series. An interrupted time series analysis of the change in GP contacts before and after introduction of UK-wide restrictions. A; grey line, the percentage of the study population with primary care contacts for each health condition. Blue region, predicted percentage of contacts from the full ITS model. Black vertical lines, data excluded data from the analysis from 1^st^ March to 29^th^ March (adjustment-to-restrictions period). B; 95% CI for the estimated relative reduction in the percentage of contacts for each health condition immediately after the adjustment-to-restrictions period (29^th^ March 2020) compared to the pre-restriction period (OR closer to 0 shows a greater reduction in the estimated percentage of people with GP contacts). C; estimated effect of time (in weekly increments) post-restriction introduction on the percentage of contacts for each condition. (OR greater that 1 indicates increasing percentage of population with contacts over time). Results for 2020 only are shown here (Figure S13 for full model fit to data from 2017) (full relative reduction/recovery estimates [95% CIs] Tables S6 and S7).

**Table 3** illustrates the potential impact of reduced contacts on relevant populations. While for some rare conditions, the absolute change in contacts was relatively small, other more common conditions had a larger absolute change in contacts, for example, in a population of 1 million people with COPD, from the end of March to the end of June, we estimated that there were cumulatively 43,900 fewer COPD exacerbation contacts per million people with COPD than expected. During the weeks commencing 26^th^ April and 28^th^ June we estimated there were 3,640 and 3,230 fewer contacts (per million) than expected respectively, indicating a slow return to pre-lockdown contact levels, but not complete recovery. For example, we estimated there were cumulatively fewer contacts for: 1) asthma exacerbations (14,100 for every one million people with asthma; and 2) depression and anxiety contacts (12,800 and 6,600 respectively) for every one million people in the general population (aged ≥11 years).

**Table 3.** Estimated reduction in estimated number of primary care contacts. Estimated number of primary care contacts for acute physical and mental health conditions in a hypothetical non-COVID year compared to the number of contacts estimated from our model for 2020 at two weekly time points: 26^th^ April and 28^th^ June 2020. Estimates for the number of contacts are in a hypothetical population of one million people but the reference populations are condition specific. For example, the estimated number of contacts for diabetic emergencies is among a population of 1 million people aged 11 years and older with a diabetes mellitus diagnosis, whereas the estimated number of people with depression contacts among one million individuals from the general population aged 5 and over.

| Condition | Week commencing | Population (age limit) | Without COVID-19 & restrictions:<br>Estimated number of contacts in a population of 1 million (95% CI) | With COVID-19 & restrictions:<br>Estimated number of contacts in a population of 1 million (95% CI) | Difference between the estimated weekly contacts and estimated number of contacts with no restrictions* | Cumulative sum of the difference in primary care contacts since 29<sup>th</sup> March* |
| --- | --- | --- | --- | --- | --- | --- |
| Diabetic emergencies | 26-Apr | People with diabetes diagnosis (11+) | 39 (34 - 44) | 14 (10 - 20) | <100 | <100 |
|  | 28-Jun | People with diabetes diagnosis (11+) | 38 (33 - 43) | 12 (8 - 19) | <100 | 330 |
| Acute alcohol-related event | 26-Apr | General population (18+) | 13 (11 - 14) | 16 (15 - 18) | >-10 | >-100 |
|  | 28-Jun | General population (18+) | 14 (13 - 16) | 24 (21 - 26) | >-10 | >-100 |
| Anxiety | 26-Apr | General population (11+) | 1,816 (1,695 - 1,945) | 1,266 (1,148 - 1,396) | 550 | 2,300 |
|  | 28-Jun | General population (11+) | 1,943 (1,818 - 2,076) | 1,532 (1,383 - 1,696) | 411 | 6,600 |
| Depression | 26-Apr | General population (11+) | 2,451 (2,285 - 2,629) | 1,391 (1,241 - 1,558) | 1,060 | 4,440 |
|  | 28-Jun | General population (11+) | 2,657 (2,484 - 2,843) | 1,857 (1,657 - 2,080) | 801 | 12,800 |
| Eating disorders | 26-Apr | General population (11+) | 44 (41 - 47) | 29 (26 - 33) | <100 | <100 |
|  | 28-Jun | General population (11+) | 47 (44 - 51) | 35 (31 - 39) | <100 | 184 |
| Obsessive compulsive disorder | 26-Apr | General population (11+) | 29 (27 - 31) | 22 (19 - 24) | <10 | <100 |
|  | 28-Jun | General population (11+) | 30 (28 - 33) | 25 (23 - 29) | <10 | <100 |
| Self-harm | 26-Apr | General population (11+) | 217 (190 - 247) | 145 (130 - 162) | <100 | 307 |
|  | 28-Jun | General population (11+) | 254 (226 - 285) | 205 (184 - 228) | <100 | 870 |
| Severe mental illness | 26-Apr | General population (11+) | 184 (173 - 196) | 155 (142 - 169) | <100 | 119 |
|  | 28-Jun | General population (11+) | 203 (192 - 215) | 172 (157 - 189) | <100 | 391 |
| Cerebrovascular accident | 26-Apr | General population (31+) | 88 (83 - 94) | 56 (50 - 62) | <100 | 135 |
|  | 28-Jun | General population (31+) | 100 (93 - 106) | 73 (65 - 81) | <100 | 400 |
| Transient ischaemic attack | 26-Apr | General population (31+) | 37 (35 - 40) | 26 (24 - 29) | <100 | <100 |
|  | 28-Jun | General population (31+) | 40 (38 - 43) | 31 (28 - 35) | <10 | 136 |
| Heart failure | 26-Apr | General population (31+) | 279 (264 - 295) | 181 (167 - 196) | <100 | 408 |
|  | 28-Jun | General population (31+) | 308 (292 - 324) | 223 (205 - 242) | <100 | 1,240 |
| Myocardial infarction | 26-Apr | General population (31+) | 45 (42 - 47) | 35 (33 - 38) | <10 | <100 |
|  | 28-Jun | General population (31+) | 47 (44 - 49) | 37 (34 - 41) | <10 | 123 |
| Unstable angina | 26-Apr | General population (31+) | 5 (5 - 6) | 4 (4 - 5) | <10 | <10 |
|  | 28-Jun | General population (31+) | 6 (5 - 6) | 6 (5 - 7) | <10 | <10 |
| Venous thromboembolism | 26-Apr | General population (31+) | 67 (63 - 70) | 64 (59 - 68) | <10 | <10 |
|  | 28-Jun | General population (31+) | 72 (69 - 76) | 63 (58 - 68) | <10 | <100 |
| Asthma exacerbations | 26-Apr | People with asthma diagnosis (11+) | 4,636 (4,361 - 4,928) | 3,617 (3,320 - 3,941) | 1,020 | 3,780 |
|  | 28-Jun | People with asthma diagnosis (11+) | 4,254 (3,995 - 4,529) | 2,941 (2,643 - 3,273) | 1,310 | 14,100 |
| COPD exacerbations | 26-Apr | People with COPD diagnosis (41+) | 7,863 (7,365 - 8,395) | 4,222 (3,768 - 4,730) | 3,640 | 14,400 |
|  | 28-Jun | People with COPD diagnosis (41+) | 6,594 (6,147 - 7,073) | 3,367 (2,919 - 3,884) | 3,230 | 43,900 |

\* Estimates of the difference between scenarios and cumulative difference have been rounded to 3 significant digits to avoid overly precise estimates. We did not intend to estimate the exact number of “missed” consultations but obtained an estimate of the absolute indirect effect of COVID-19 on different conditions. If the expected difference was <100 or <10 then estimates have been censored for the same reason.
Abbreviations: CI: confidence interval. COPD: chronic obstructive pulmonary disease

## DISCUSSION

Primary care contacts for key physical and mental health conditions dropped dramatically after the introduction of population-wide restriction measures in March 2020. By July 2020, with the exception of unstable angina and acute alcohol-related contacts, primary care contacts for all conditions studied remained below pre-lockdown levels. We estimate that by July 2020, per million people in the general population, there were very small (<10) drops in contacts for myocardial infarction, unstable angina and venous thromboembolism, but drops of around 6,600 and 12,800 anxiety and depression contacts. Per million people with COPD, we estimate 43,900 fewer exacerbation contacts.

### Results in context

Our study is the first to explore the effect of lockdown measures on primary care contacts for specific acute physical and mental health conditions across the UK. A study (47 primary care practices) in a largely deprived urban area, badly affected by the pandemic in North West England (Salford) suggested primary care consultations across four broad categories (common mental health problems, cardiovascular and cerebrovascular disease, type 2 diabetes, and cancer) had reduced by up to 50% by the end of May 2020.^18^ In contrast to the Salford study, our sample was nationally representative and focused on contacts for specific disease categories we would expect to present to healthcare providers. Our large sample size allowed us to investigate detailed diagnoses (for example, different types of CVD and mental health conditions).

In September 2020 GPs conducted more face-to-face appointments than any week since March, and more consultations overall than prior to the pandemic (43% were telephone appointments).^30,31^ A study of 51 GP practices already offering remote consultations before the pandemic indicated a dip in overall consultations at lockdown, but unlike our results for specific acute conditions, their post-lockdown overall consultation decrease was less extreme than during Christmas 2019.^32^ In England there was a 30% decrease in GP consultations from the beginning to the end of March 2020,^33^ with an increase in calls to NHS 111, the non-urgent telephone helpline. However, over 50% (1,573,835/2,962,751) of these calls went unanswered.^34^

#### Diabetic emergencies

The reduced diabetic emergency contacts we observed are consistent with the 49% reduction in new type 2 diabetes contacts (new prescriptions for metformin) in Salford. While the Salford study highlighted missed new diagnoses, our study identifies missed contacts for acute deteriorations. Given that 90% of diabetes management is in primary care, the large relative reduction in the proportion of people with diabetes with diabetic emergency contacts is concerning.^35^

Recent evidence indicates a two-way interaction between diabetes and COVID-19, with a potentially causal association between COVID-19 infection and dysglycaemia, such that each condition exacerbates the other.^36,37^ Further, there is evidence that other emergency situations impair control of diabetes.^38–40^ Consequently, we would expect an increase, rather than decrease, in diabetic emergency contacts. One potential explanation for our findings is that individuals experiencing diabetic emergencies may be presenting directly to secondary care with delayed recording in the primary care health record.

### Cardiovascular disease

The reduction in CVD contacts is consistent with reports from other UK studies.^18,41^ Taken alongside findings of similar reductions in Emergency Department presentations and hospital admissions for cardiovascular outcomes in the UK, our findings highlight an area of major concern,^3,42^ particularly as evidence from France indicates increased out-of-hospital cardiac arrest.^43^ Severe COVID-19 affects the cardiovascular system;^44^ hence, increased primary and secondary care presentations for CVD are expected.^45^ Indeed, it is possible that the more rapid recovery in unstable angina contacts (compared to other study conditions) may reflect COVID-19-related CVD (however, numbers were small).^46^

### Respiratory disease

Reports from Germany, consistent with our findings, indicate reduced community and hospital presentations with acute COPD exacerbations.^47^ COPD is associated with more severe COVID-19 infection;^48^ individuals with COPD in the UK were recommended to avoid contact with others until September 2020.^19,49^ Decreased emergency department visits for childhood asthma have been reported in the USA, consistent with our observations.^50^ There is no compelling evidence that individuals with asthma are at greater risk of severe COVID-19 outcomes, although there was uncertainty at pandemic onset.^51–53^ Viruses commonly trigger asthma exacerbations, so we might have expected to see more asthma contacts. Anecdotally, GPs reported increased prescription of asthma therapies around the lockdown period,^54^ which may explain initial increased asthma contacts. Similar increases in COPD exacerbation contacts were not seen around introduction of restrictions, despite our definition including prescriptions for oral corticosteroids. One explanation might be that, as COPD is a progressive respiratory condition, individuals with COPD may have repeat prescriptions, reducing need (compared to those with asthma) to stockpile drugs in a crisis.

### Mental health

Surveys have reported increased anxiety, depression and self-harm during the pandemic,^12,13,55–57^ and exacerbations of existing OCD, severe mental illness (SMI) and eating disorders have also been reported.^58–60^ However, we saw a sustained reduction in primary care contacts for anxiety, depression and other mental health conditions consistent with other reports;^18^ this is concerning as the majority of common mental disorders are managed in primary care. Similarly, reduced healthcare contacts for people with SMIs are concerning, as people with SMIs are likely to be at greater risk of poor outcomes from COVID-19 due to high prevalence of risk factors for adverse outcomes (e.g. CVD, deprivation^61–63^).

### Alcohol

Findings from surveys on alcohol consumption in lockdown are mixed, with some reporting increased alcohol consumption in up to a third of those surveyed, while others produced differing findings.^64^ We saw primary care contacts for acute alcohol-related events increase before and after restrictions, which is troubling given the reduction in contacts for other conditions studied (however, numbers were small).

## Strengths and limitations

We performed a rapid assessment of changes in primary care contacts following the introduction of UK-population-wide restrictions up to July 2020 in a large sample respresentative of the UK population. Historical data allowed us to compare observed patterns in 2020 to trends in the previous 3-years. We estimated relative and absolute changes in contact patterns, with a focus on easy to interpret measures.

Our study describes and quantifies the reduction in primary care contacts across a wide range of health conditions likely to be affected by COVID-19 to generate hypotheses. Further research is needed to understand the specific drivers behind these changes. It is important that we understand what happened to individuals who did not consult their GP. Specifically, were they treated in secondary care or did they self-manage, and how much of our findings can be explained by genuine changes in disease frequency?

Without hospital and mortality data, we are unable to investigate whether, for example, any reduction in GP contacts resulted in corresponding increases in hospital attendances or deaths. We focused on studying any record of study conditions, hence, our results reflect all primary care contacts, including diagnoses recorded by general practice staff from hospital discharge letters.

To avoid problems arising from the timing of behaviour change associated with restrictions, our ITS analysis excluded a predefined intervention period when individuals’ behaviour was changing dynamically. We took a conservative approach and defined our intervention period between 1^st^ March and 28^th^ March 2020 assuming that some people would have modified behaviour before the introduction of restrictions. Sensitivity analyses varying the start date showed consistent findings.

Detailed exploration of whether consultation behaviour varied in those considered clinically vulnerable and advised to ‘shield^18^ is beyond the scope of this paper, and any changes in health-seeking behaviour would not have reduced the need for care.

## Clinical interpretation

Given evidence suggesting reduced emergency department attendances and hospital admissions for our study conditions, while one explanation could be genuine changes in disease frequency (unlikely, given consistent results across disease categories), it is more likely our findings reflect missed opportunities for care. There are plausible mechanisms that might explain real reductions in frequency for some of our outcomes. For example: 1) better glycaemic control in diabetes due to more regular routines when staying home; 2) less respiratory disease due to lower exposure to air pollution during lockdown,^65^ and reduced community-aquired respiratory infections due to ‘shielding’ guidelines;^19^ and 3) reduced alcohol consumption due to pub closure and reduced social contact. Conversely, there are plausible mechanisms that could explain genuine increased frequency of these conditions (e.g. distress related to the pandemic affecting mental health and alcohol consumption, reduced exercise affecting cardiovascular health, changes in diet influencing glycaemic control); and for some of our outcomes, e.g., mental health conditions, there is some evidence indicating increased frequency.^12,13,55,56,58–60^ Increases in non-COVID-related excess mortality also make it more likely our observed reduction in primary care contacts was due to behavioural changes rather than reduced disease frequency.^13,66–69^ Further, emerging evidence of the systemic complications of COVID-19 infection (particularly CVD and diabetes)^36,70,71^ indicates we might have expected more need for care for these conditions as a direct result of the pandemic.

## Implications for policy and research

Our results are likely to represent a large burden of unmet need, particularly in relation to COPD and mental health conditions; healthcare providers should prepare for increases in morbidity and mortality. Further research should address whether reduced clinical contact has resulted in excess mortality, and whether we need to increase service provision for individuals with increased healthcare needs resulting from delaying seeking access to care. While numbers of unstable angina events were small, we note more rapid (compared to other study outcomes) return to pre-pandemic consultation rates; this observation needs investigation as it may be a direct consequence of the pandemic. Finally, our findings highlight a need to ensure equitable access to primary care in future pandemic planning. Countries such as Singapore, which had experienced SARS, implemented control measures in primary care rapidly.^72^ The current pandemic has generated a wealth of experience with alternative ways to access care remotely.^73^ These lessons must be systematised and implemented.

## Conclusions

We saw substantial reductions in primary care contacts for acute physical and mental health conditions. Our findings are likely to represent a considerable burden of unmet need, which may lead to substantial increases in subsequent mortality and morbidity.

## Data Availability

No additional unpublished data are available as this study used existing data from the UK CPRD electronic health record database that is only accessible to researchers with protocols approved by the CPRD's Independent Scientific Advisory Committee.
All data management and analysis computer code is available via GitHub (https://github.com/johntaz/COVID-Collateral). All code is shared without investigator support. Our study protocol and analysis plan are available as additional online-only supplementary material. All aggregated data will be freely available to explore by stratifiers through an R Shiny app, available at https://a-henderson91.shinyapps.io/covid_collateral_shiny/.

https://github.com/johntaz/COVID-Collateral

https://a-henderson91.shinyapps.io/covid_collateral_shiny/

## DECLARATIONS AND ACKNOWLEDGEMENTS

### Contributions

All study authors were involved in the development of the study. KEM, RM, JT, AH and AM contributed equally and are considered joint first authors. All authors contributed to the development of the code lists that defined the variables used in the study. JT, AH, RM, PB, HC, and AW were responsible for data management. JT, AH, RM and AM were responsible for statistical analyses. KEM, RM, JT, AH, and AM wrote the first paper draft. All authors contributed to and approved the final manuscript.

## Acknowledgements

We would like to thank John Burn-Murdoch, as our main plot design was based on his work on excess mortality in the Financial Times (https://www.ft.com/content/a2901ce8-5eb7-4633-b89c-cbdf5b386938).

This study is based in part on data from the Clinical Practice Research Datalink obtained under licence from the UK Medicines and Healthcare products Regulatory Agency. The data is provided by patients and collected by the NHS as part of their care and support. The interpretation and conclusions contained in this study are those of the authors alone. The study was approved by the Independent Scientific Advisory Committee (Protocol number: 20_089R2).

## Ethical approval

The study was approved by the London School of Hygiene and Tropical Medicine Research Ethics Committee (Reference: 22143 /RR/18495) and by the CPRD Independent Scientific Advisory Committee (ISAC Protocol Number: 20_089R2).

Data sharing

No additional unpublished data are available as this study used existing data from the UK CPRD electronic health record database that is only accessible to researchers with protocols approved by the CPRD’s Independent Scientific Advisory Committee.

All data management and analysis computer code is available via GitHub (https://github.com/johntaz/COVID-Collateral). All code is shared without investigator support. Our study protocol and analysis plan are available as additional online-only supplementary material. All aggregated data will be freely available to explore by stratifiers through an R Shiny app, available at https://a-henderson91.shinyapps.io/covid_collateral_shiny/.

