## Supplementary material for "COVID-19 collateral: Indirect acute effects of the pandemic on physical and mental health in the UK"

*Figure S1. Illustration of overall study population and condition-specific study populations.*

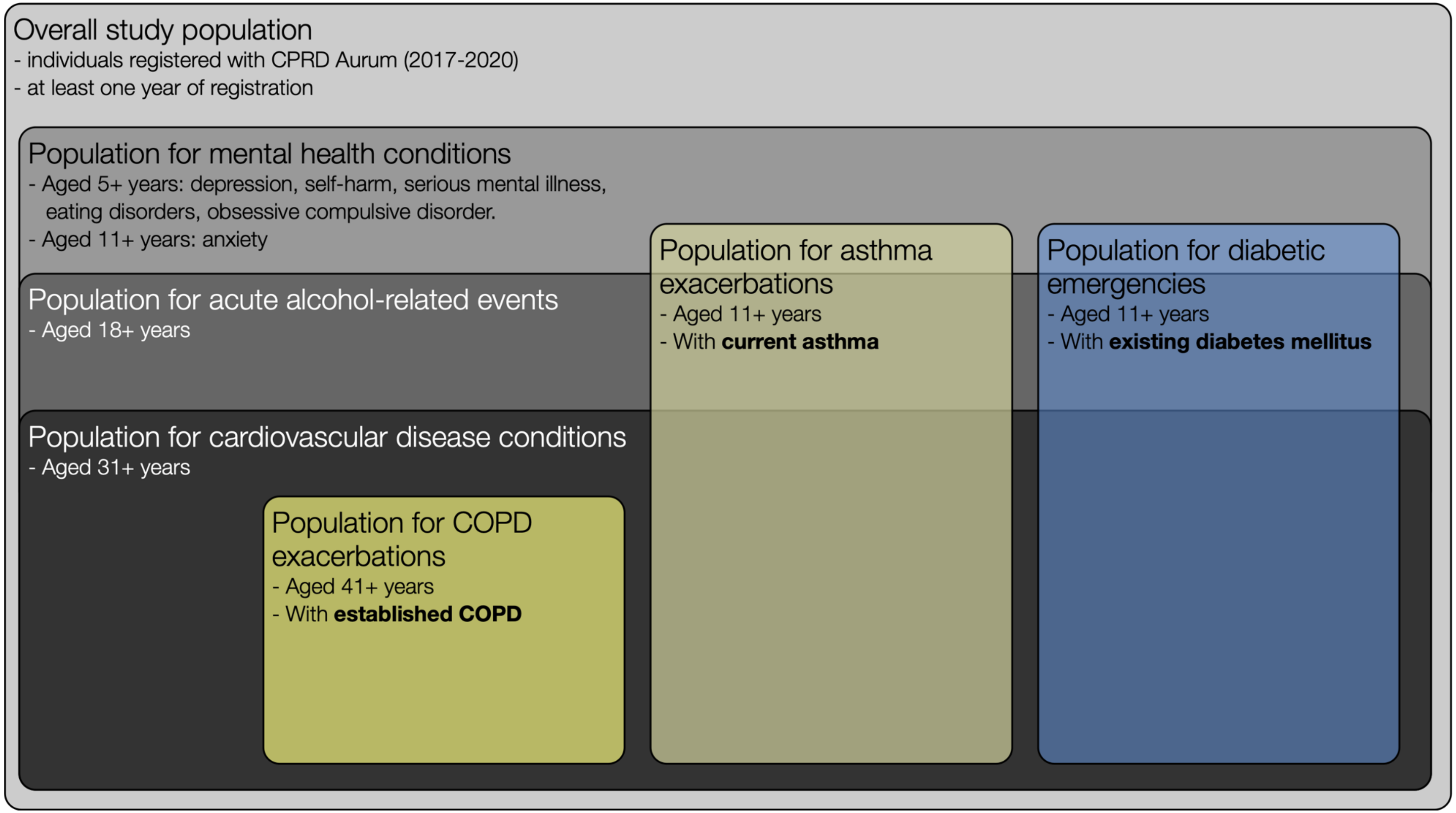

NB: Relative size of boxes do not represent relative sizes of study populations. For simplicity we have not shown the boxes for the study populations with asthma, COPD and diabetes overlapping, However, it is likely that there will be some individuals with multiple morbidities included in multiple populations, for example, there will be some people with both diabetes and COPD.

**Abbreviations**: COPD, chronic obstructive pulmonary disease; CPRD, Clinical Practice Research Datalink

### Condition-specific denominator populations

*Table S1 – Description of the denominator population for acute alcohol-related events, as measured in the first week of January 2017-2020. Percentages of total denominator population are shown in parentheses.*

| | Category | 2017 | | 2018 | | 2019 | | 2020 | |
|---|---|---|---|---|---|---|---|---|---|
| | Overall denominator | 8,974,499 | (100) | 9,195,503 | (100) | 9,329,369 | (100) | 9,264,471 | (100) |
| Age (years) | 18 - 20 | 343,983 | (4) | 354,773 | (4) | 362,880 | (4) | 362,944 | (4) |
| | 21 - 30 | 1,455,550 | (16) | 1,499,066 | (16) | 1,517,439 | (16) | 1,505,172 | (16) |
| | 31 - 40 | 1,559,933 | (17) | 1,622,838 | (18) | 1,662,883 | (18) | 1,661,724 | (18) |
| | 41 - 50 | 1,577,507 | (18) | 1,579,296 | (17) | 1,573,889 | (17) | 1,550,104 | (17) |
| | 51 - 60 | 1,520,720 | (17) | 1,564,290 | (17) | 1,590,738 | (17) | 1,580,348 | (17) |
| | 61 - 70 | 1,165,390 | (13) | 1,166,078 | (13) | 1,176,134 | (13) | 1,164,688 | (13) |
| | 71 - 80 | 833,570 | (9) | 881,099 | (10) | 907,289 | (10) | 904,486 | (10) |
| | 81 - 90 | 426,769 | (5) | 436,646 | (5) | 445,112 | (5) | 442,098 | (5) |
| | 91 - 100 | 91,077 | (1) | 91,417 | (1) | 93,005 | (1) | 92,907 | (1) |
| Ethnicity | White | 4,513,594 | (50) | 4,624,221 | (50) | 4,697,089 | (50) | 4,595,278 | (50) |
| | South Asian | 383,479 | (4) | 404,874 | (4) | 413,318 | (4) | 426,361 | (5) |
| | Black | 232,746 | (3) | 242,300 | (3) | 243,346 | (3) | 248,200 | (3) |
| | Other | 136,663 | (2) | 150,460 | (2) | 163,184 | (2) | 173,055 | (2) |
| | Mixed | 80,430 | (1) | 86,359 | (1) | 90,944 | (1) | 94,377 | (1) |
| | Missing | 3,627,587 | (40) | 3,687,289 | (40) | 3,721,488 | (40) | 3,727,200 | (40) |
| Sex | Female | 4,489,298 | (50) | 4,593,678 | (50) | 4,658,434 | (50) | 4,621,873 | (50) |
| | Male | 4,485,201 | (50) | 4,601,825 | (50) | 4,670,935 | (50) | 4,642,598 | (50) |
| Region | North East | 315,629 | (4) | 319,354 | (4) | 323,973 | (4) | 313,763 | (3) |
| | North West | 1,539,272 | (17) | 1,567,260 | (17) | 1,592,118 | (17) | 1,602,856 | (17) |
| | Yorkshire and the Humber | 341,192 | (4) | 349,911 | (4) | 357,458 | (4) | 329,935 | (4) |
| | East Midlands | 238,915 | (3) | 246,463 | (3) | 255,619 | (3) | 214,920 | (2) |
| | West Midlands | 1,427,095 | (16) | 1,452,751 | (16) | 1,450,408 | (16) | 1,469,456 | (16) |
| | East of England | 419,707 | (5) | 425,830 | (5) | 424,747 | (5) | 388,164 | (4) |
| | South West | 1,083,665 | (12) | 1,110,448 | (12) | 1,110,511 | (12) | 1,097,636 | (12) |
| | South Central | 1,126,410 | (13) | 1,150,518 | (13) | 1,164,597 | (13) | 1,175,184 | (13) |
| | London | 1,672,917 | (19) | 1,749,909 | (19) | 1,806,520 | (19) | 1,833,233 | (20) |
| | South East Coast | 752,133 | (8) | 764,640 | (8) | 785,323 | (8) | 780,288 | (8) |
| | Northern Ireland | 41,268 | (1) | 42,127 | (1) | 42,983 | (1) | 43,913 | (1) |

*Table S2 – Description of the denominator population for asthma exacerbations, as measured in the first week of January 2017-2020. Percentages of total denominator population are shown in parentheses.*

| | **Category** | **2017** | | **2018** | | **2019** | | **2020** | |
|---|---|---|---|---|---|---|---|---|---|
| **Overall** | Overall denominator | 882,141 | (100) | 911,579 | (100) | 930,478 | (100) | 925,795 | (100) |
| **Age (years)** | 11 - 20 | 126,046 | (14) | 131,451 | (14) | 134,201 | (14) | 132,958 | (14) |
| | 21 - 30 | 121,166 | (14) | 126,651 | (14) | 130,488 | (14) | 131,277 | (14) |
| | 31 - 40 | 130,758 | (15) | 135,257 | (15) | 137,348 | (15) | 135,432 | (15) |
| | 41 - 50 | 151,529 | (17) | 151,355 | (17) | 150,289 | (16) | 146,610 | (16) |
| | 51 - 60 | 142,438 | (16) | 148,077 | (16) | 151,898 | (16) | 151,681 | (16) |
| | 61 - 70 | 102,330 | (12) | 103,790 | (11) | 105,957 | (11) | 106,028 | (12) |
| | 71 - 80 | 69,801 | (8) | 74,576 | (8) | 77,525 | (8) | 78,023 | (8) |
| | 81 - 90 | 32,877 | (4) | 34,870 | (4) | 36,675 | (4) | 37,367 | (4) |
| | 91 - 100 | 5,196 | (1) | 5,552 | (1) | 6,097 | (1) | 6,419 | (1) |
| **Ethnicity** | White | 485,515 | (55) | 502,818 | (55) | 515,848 | (55) | 509,045 | (55) |
| | South Asian | 41,203 | (5) | 43,692 | (5) | 44,426 | (5) | 45,405 | (5) |
| | Black | 21,876 | (3) | 23,040 | (3) | 23,424 | (3) | 23,991 | (3) |
| | Other | 6,956 | (1) | 7,623 | (1) | 8,132 | (1) | 8,533 | (1) |
| | Mixed | 9,498 | (1) | 10,434 | (1) | 11,028 | (1) | 11,477 | (1) |
| | Missing | 317,093 | (36) | 323,972 | (36) | 327,620 | (35) | 327,344 | (35) |
| **Sex** | Female | 500,972 | (57) | 514,796 | (57) | 522,970 | (56) | 517,863 | (56) |
| | Male | 381,169 | (43) | 396,783 | (44) | 407,508 | (44) | 407,932 | (44) |
| **Region** | North East | 30,297 | (3) | 31,003 | (3) | 31,760 | (3) | 31,015 | (3) |
| | North West | 158,004 | (18) | 162,735 | (18) | 166,655 | (18) | 168,060 | (18) |
| | Yorkshire and the Humber | 32,373 | (4) | 33,480 | (4) | 34,379 | (4) | 31,978 | (4) |
| | East Midlands | 23,406 | (3) | 24,587 | (3) | 25,410 | (3) | 21,377 | (2) |
| | West Midlands | 144,115 | (16) | 148,309 | (16) | 149,439 | (16) | 152,152 | (16) |
| | East of England | 43,360 | (5) | 44,275 | (5) | 44,333 | (5) | 40,822 | (4) |
| | South West | 112,592 | (13) | 116,876 | (13) | 118,531 | (13) | 117,871 | (13) |
| | South Central | 115,881 | (13) | 119,356 | (13) | 121,633 | (13) | 122,852 | (13) |
| | London | 144,994 | (16) | 151,694 | (17) | 156,521 | (17) | 158,105 | (17) |
| | South East Coast | 71,168 | (8) | 73,186 | (8) | 75,748 | (8) | 75,400 | (8) |
| | Northern Ireland | 4,429 | (1) | 4,536 | (1) | 4,627 | (1) | 4,709 | (1) |

*Table S3 – Description of the denominator population for COPD exacerbations, as measured in the first week of January 2017-2020. Percentages of total denominator population are shown in parentheses.*

| | Category | 2017 | | 2018 | | 2019 | | 2020 | |
|---|---|---|---|---|---|---|---|---|---|
| **Overall** | Overall denominator | 283,406 | (100) | 272,201 | (100) | 259,039 | (100) | 239,809 | (100) |
| **Age (years)** | 11 - 20 | - | - | - | - | - | - | - | - |
| | 21 - 30 | - | - | - | - | - | - | - | - |
| | 31 - 40 | - | - | - | - | - | - | - | - |
| | 41 - 50 | 14,445 | (5) | 12,271 | (5) | 10,312 | (4) | 8,349 | (4) |
| | 51 - 60 | 46,909 | (17) | 43,846 | (16) | 40,586 | (16) | 36,369 | (15) |
| | 61 - 70 | 83,332 | (29) | 77,256 | (28) | 71,914 | (28) | 65,626 | (27) |
| | 71 - 80 | 88,906 | (31) | 88,889 | (33) | 86,442 | (33) | 81,384 | (34) |
| | 81 - 90 | 43,636 | (15) | 43,637 | (16) | 43,304 | (17) | 41,648 | (17) |
| | 91 - 100 | 6,178 | (2) | 6,302 | (2) | 6,481 | (3) | 6,433 | (3) |
| **Ethnicity** | White | 182,453 | (64) | 175,964 | (65) | 167,920 | (65) | 154,549 | (64) |
| | South Asian | 3,048 | (1) | 3,037 | (1) | 2,942 | (1) | 2,851 | (1) |
| | Black | 2,086 | (1) | 2,033 | (1) | 1,915 | (1) | 1,843 | (1) |
| | Other | 766 | (0) | 759 | (0) | 754 | (0) | 731 | (0) |
| | Mixed | 767 | (0) | 767 | (0) | 726 | (0) | 689 | (0) |
| | Missing | 94,286 | (33) | 89,641 | (33) | 84,782 | (33) | 79,146 | (33) |
| **Sex** | Female | 132,967 | (47) | 128,166 | (47) | 122,419 | (47) | 113,825 | (48) |
| | Male | 150,439 | (53) | 144,035 | (53) | 136,620 | (53) | 125,984 | (53) |
| **Region** | North East | 14,877 | (5) | 14,154 | (5) | 13,498 | (5) | 12,298 | (5) |
| | North West | 60,924 | (22) | 58,435 | (22) | 55,797 | (22) | 52,519 | (22) |
| | Yorkshire and the Humber | 11,409 | (4) | 11,025 | (4) | 10,515 | (4) | 9,042 | (4) |
| | East Midlands | 6,905 | (2) | 6,601 | (2) | 6,353 | (3) | 4,717 | (2) |
| | West Midlands | 47,419 | (17) | 45,777 | (17) | 43,713 | (17) | 41,547 | (17) |
| | East of England | 12,366 | (4) | 11,776 | (4) | 10,897 | (4) | 9,247 | (4) |
| | South West | 37,036 | (13) | 35,626 | (13) | 33,180 | (13) | 30,647 | (13) |
| | South Central | 30,381 | (11) | 29,035 | (11) | 27,586 | (11) | 26,097 | (11) |
| | London | 36,756 | (13) | 35,505 | (13) | 34,057 | (13) | 31,941 | (13) |
| | South East Coast | 23,229 | (8) | 22,228 | (8) | 21,479 | (8) | 19,876 | (8) |
| | Northern Ireland | 1,585 | (1) | 1,538 | (1) | 1,492 | (1) | 1,428 | (1) |

*Table S4 – Description of the denominator population for cardiovascular conditions, as measured in the first week of January 2017-2020. Percentages of total denominator population are shown in parentheses.*

| | Category | 2017 | | 2018 | | 2019 | | 2020 | |
|---|---|---|---|---|---|---|---|---|---|
| **Overall** | Overall denominator | 7,174,966 | (100) | 7,341,664 | (100) | 7,449,050 | (100) | 7,396,355 | (100) |
| **Age** | 11 - 20 | - | - | - | - | - | - | - | - |
| | 21 - 30 | - | - | - | - | - | - | - | - |
| | 31 - 40 | 1,559,933 | (22) | 1,622,838 | (22) | 1,662,883 | (22) | 1,661,724 | (23) |
| | 41 - 50 | 1,577,507 | (22) | 1,579,296 | (22) | 1,573,889 | (21) | 1,550,104 | (21) |
| | 51 - 60 | 1,520,720 | (21) | 1,564,290 | (21) | 1,590,738 | (21) | 1,580,348 | (21) |
| | 61 - 70 | 1,165,390 | (16) | 1,166,078 | (16) | 1,176,134 | (16) | 1,164,688 | (16) |
| | 71 - 80 | 833,570 | (12) | 881,099 | (12) | 907,289 | (12) | 904,486 | (12) |
| | 81 - 90 | 426,769 | (6) | 436,646 | (6) | 445,112 | (6) | 442,098 | (6) |
| | 91 - 100 | 91,077 | (1) | 91,417 | (1) | 93,005 | (1) | 92,907 | (1) |
| **Ethnicity** | White | 3,779,955 | (53) | 3,871,985 | (53) | 3,937,363 | (53) | 3,858,594 | (52) |
| | South Asian | 293,049 | (4) | 312,242 | (4) | 320,937 | (4) | 332,222 | (5) |
| | Black | 180,227 | (3) | 187,706 | (3) | 188,628 | (3) | 192,517 | (3) |
| | Other | 86,473 | (1) | 94,994 | (1) | 102,469 | (1) | 109,191 | (2) |
| | Mixed | 56,005 | (1) | 60,115 | (1) | 63,305 | (1) | 65,851 | (1) |
| | Missing | 2,779,257 | (39) | 2,814,622 | (38) | 2,836,348 | (38) | 2,837,980 | (38) |
| **Sex** | Female | 3,618,289 | (50) | 3,695,459 | (50) | 3,745,435 | (50) | 3,712,398 | (50) |
| | Male | 3,556,677 | (50) | 3,646,205 | (50) | 3,703,615 | (50) | 3,683,957 | (50) |
| **Region** | North East | 248,280 | (4) | 250,999 | (3) | 254,945 | (3) | 246,165 | (3) |
| | North West | 1,225,731 | (17) | 1,245,139 | (17) | 1,263,473 | (17) | 1,272,387 | (17) |
| | Yorkshire and the Humber | 259,709 | (4) | 265,218 | (4) | 269,219 | (4) | 245,151 | (3) |
| | East Midlands | 172,559 | (2) | 178,118 | (2) | 183,218 | (3) | 147,195 | (2) |
| | West Midlands | 1,157,537 | (16) | 1,177,622 | (16) | 1,178,910 | (16) | 1,195,451 | (16) |
| | East of England | 354,740 | (5) | 359,343 | (5) | 358,035 | (5) | 326,440 | (4) |
| | South West | 878,048 | (12) | 896,719 | (12) | 895,673 | (12) | 883,609 | (12) |
| | South Central | 910,667 | (13) | 928,185 | (13) | 939,790 | (13) | 950,188 | (13) |
| | London | 1,309,139 | (18) | 1,370,179 | (19) | 1,419,386 | (19) | 1,447,147 | (20) |
| | South East Coast | 613,558 | (9) | 624,457 | (9) | 640,857 | (9) | 636,441 | (9) |
| | Northern Ireland | 31,721 | (0) | 32,401 | (0) | 33,069 | (0) | 33,667 | (1) |

*Table S5 – Description of the denominator population for diabetic emergencies, as measured in the first week of January 2017-2020. Percentages of total denominator population are shown in parentheses.*

| | Category | 2017 | | 2018 | | 2019 | | 2020 | |
|---|---|---|---|---|---|---|---|---|---|
| **Overall** | Overall denominator | 699,396 | (100) | 690,707 | (100) | 674,150 | (100) | 643,682 | (100) |
| **Age** | 11 - 20 | 6,009 | (1) | 5,884 | (1) | 5,671 | (1) | 5,247 | (1) |
| | 21 - 30 | 11,036 | (2) | 10,729 | (2) | 10,378 | (2) | 9,833 | (2) |
| | 31 - 40 | 28,004 | (4) | 26,651 | (4) | 24,929 | (4) | 22,850 | (4) |
| | 41 - 50 | 76,508 | (11) | 71,678 | (10) | 66,118 | (10) | 59,991 | (9) |
| | 51 - 60 | 143,037 | (21) | 139,862 | (20) | 134,782 | (20) | 127,329 | (20) |
| | 61 - 70 | 172,312 | (25) | 167,644 | (24) | 163,513 | (24) | 156,783 | (24) |
| | 71 - 80 | 160,657 | (23) | 163,421 | (24) | 162,095 | (24) | 156,375 | (24) |
| | 81 - 90 | 88,587 | (13) | 90,987 | (13) | 92,066 | (14) | 90,370 | (14) |
| | 91 - 100 | 13,246 | (2) | 13,851 | (2) | 14,598 | (2) | 14,904 | (2) |
| **Ethnicity** | White | 364,403 | (52) | 358,470 | (52) | 349,154 | (52) | 327,187 | (51) |
| | South Asian | 53,148 | (8) | 54,242 | (8) | 53,394 | (8) | 52,862 | (8) |
| | Black | 26,619 | (4) | 26,888 | (4) | 26,052 | (4) | 25,594 | (4) |
| | Other | 6,695 | (1) | 7,080 | (1) | 7,255 | (1) | 7,380 | (1) |
| | Mixed | 5,843 | (1) | 5,972 | (1) | 5,952 | (1) | 5,842 | (1) |
| | Missing | 242,688 | (35) | 238,055 | (35) | 232,343 | (35) | 224,817 | (35) |
| **Sex** | Female | 309,214 | (44) | 305,090 | (44) | 297,288 | (44) | 283,621 | (44) |
| | Male | 390,182 | (56) | 385,617 | (56) | 376,862 | (56) | 360,061 | (56) |
| **Region** | North East | 25,575 | (4) | 25,037 | (4) | 24,485 | (4) | 22,901 | (4) |
| | North West | 123,120 | (18) | 120,535 | (18) | 117,708 | (18) | 113,871 | (18) |
| | Yorkshire And The Humber | 25,341 | (4) | 25,025 | (4) | 24,454 | (4) | 21,353 | (3) |
| | East Midlands | 16,751 | (2) | 16,642 | (2) | 16,380 | (2) | 12,458 | (2) |
| | West Midlands | 124,167 | (18) | 121,841 | (18) | 116,894 | (17) | 114,183 | (18) |
| | East of England | 29,713 | (4) | 29,148 | (4) | 27,858 | (4) | 23,943 | (4) |
| | South West | 83,235 | (12) | 82,172 | (12) | 78,483 | (12) | 74,385 | (12) |
| | South Central | 78,777 | (11) | 77,553 | (11) | 75,859 | (11) | 73,981 | (12) |
| | London | 133,720 | (19) | 134,642 | (20) | 134,380 | (20) | 131,521 | (20) |
| | South East Coast | 54,599 | (8) | 53,806 | (8) | 53,501 | (8) | 51,053 | (8) |
| | Northern Ireland | 2,831 | (0) | 2,789 | (0) | 2,725 | (0) | 2,653 | (0) |

### Trends for each condition by stratification variables

#### *Figure S2 – Stratified by ethnicity*

Percentage of study populations with primary care contacts for each health condition over 2020, by ethnicity. Boxplots show the historical average percentage of study population with GP contacts for the condition of interest. Coloured lines, weekly percentage of eligible population with GP contacts for the condition of interest in 2020. Red dotted line, introduction of restrictions in UK on March 23rd 2020. If ethnicity information was missing then data are not shown. Note that cell counts with fewer than five contacts in one week in 2020 have been suppressed.

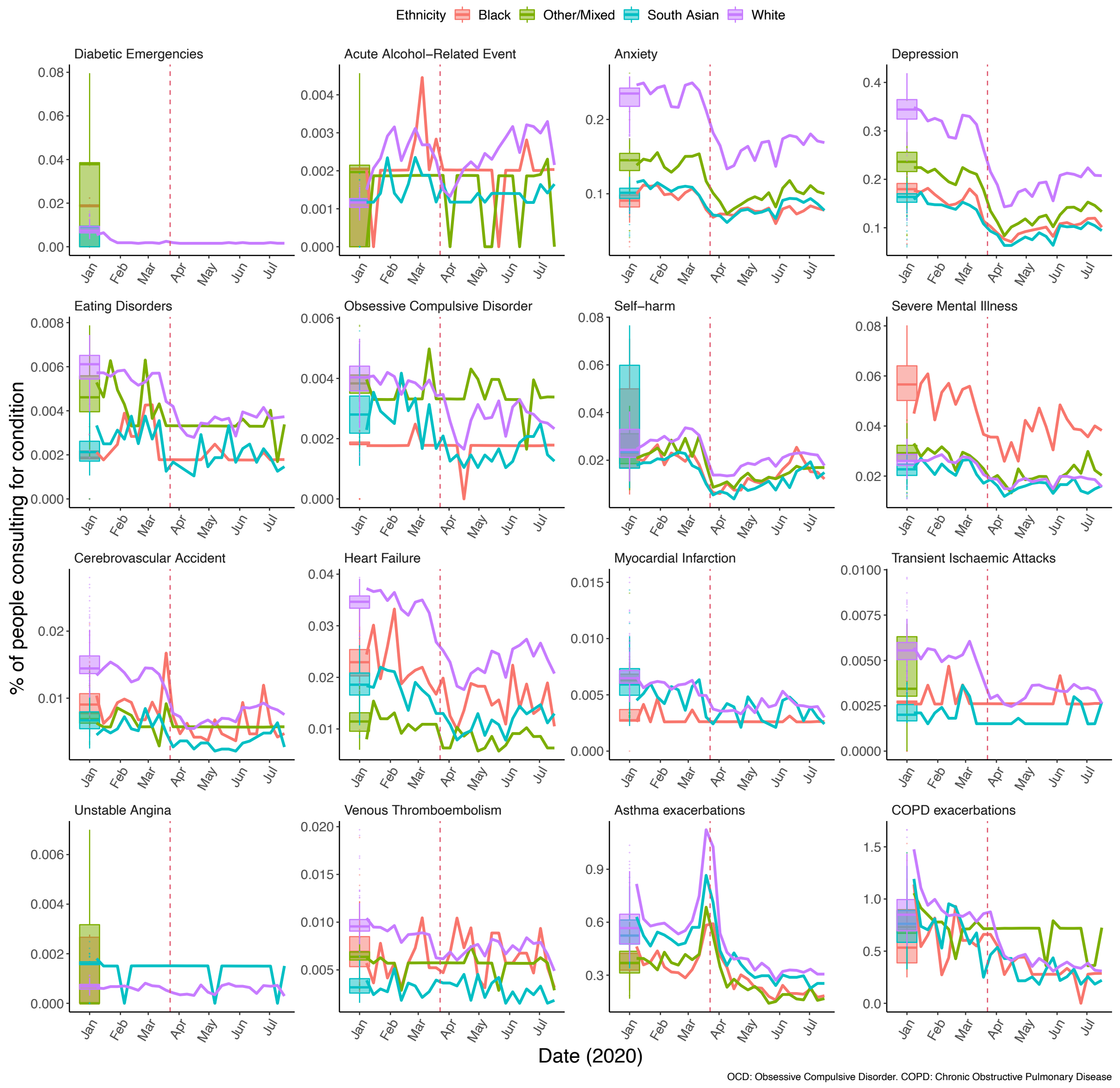

### *Figure S3 – Stratified by sex*

Percentage of study populations with primary care contacts for each health condition over 2020, by sex. Boxplots show the historical average percentage of study populations with GP contancts for the condition of interest. Coloured lines, weekly percentage of eligible population that with GP contacts for the condition of interest in 2020. Red dotted line, introduction of restrictions in UK on March 23rd 2020. Note that cell counts with fewer than 5 outcomes in one week in 2020 have been suppressed.

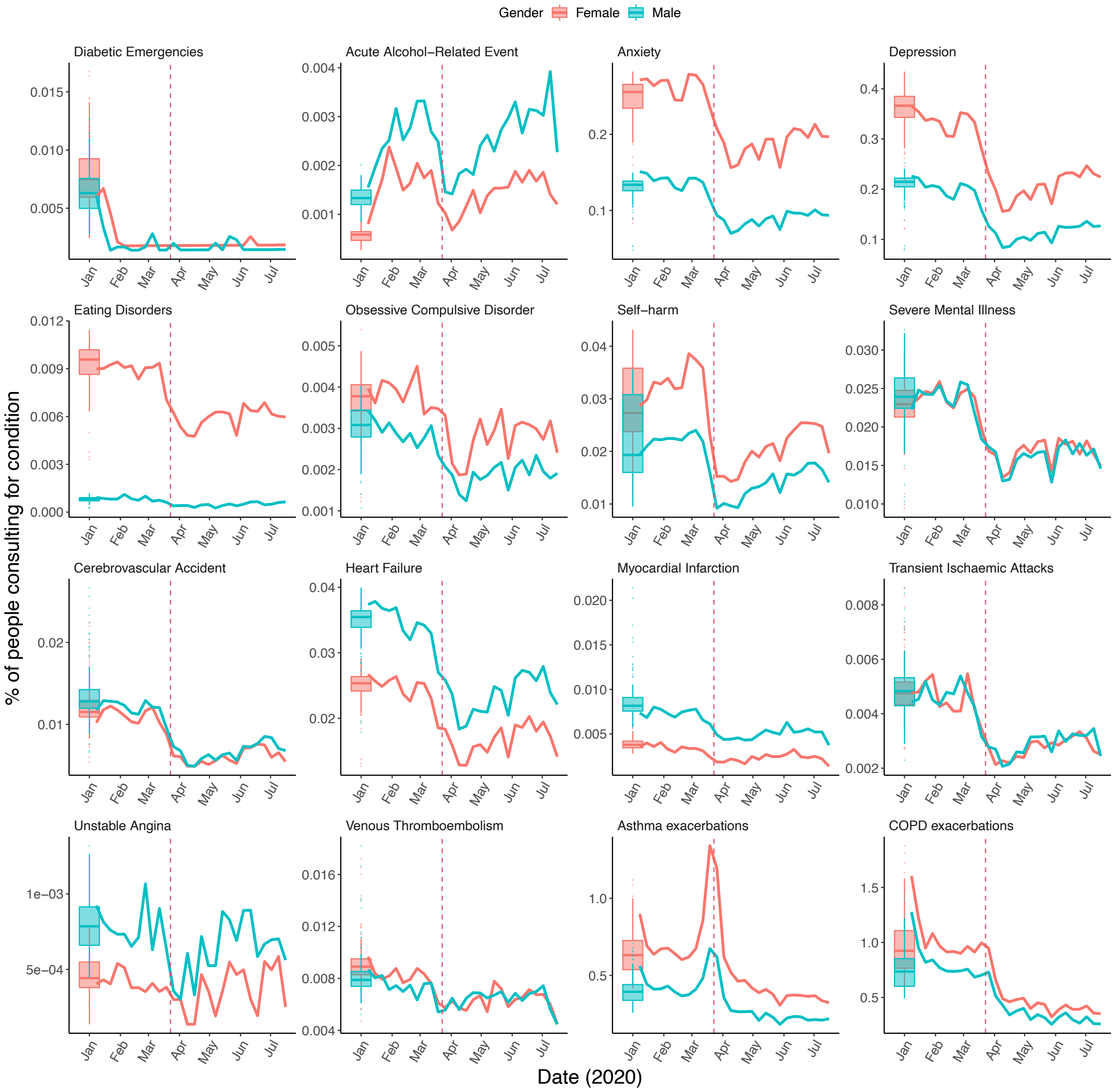

OCD: Obsessive Compulsive Disorder. COPD: Chronic Obstructive Pulmonary Disease

***Figure S4 – Stratified by geographic region.***

Percentage of study populations with GP contacts for each health condition over 2020, by region. Boxplots show the historical average percentage of study populations with GP contacts for the condition of interest. Coloured lines, weekly percentage of eligible population with GP contacts for the condition of interest in 2020. Red dotted line, introduction of restrictions in UK on March 23rd 2020. Data are not shown if information on region was missing and cell counts with fewer than five contacts in one week in 2020 have been suppressed.

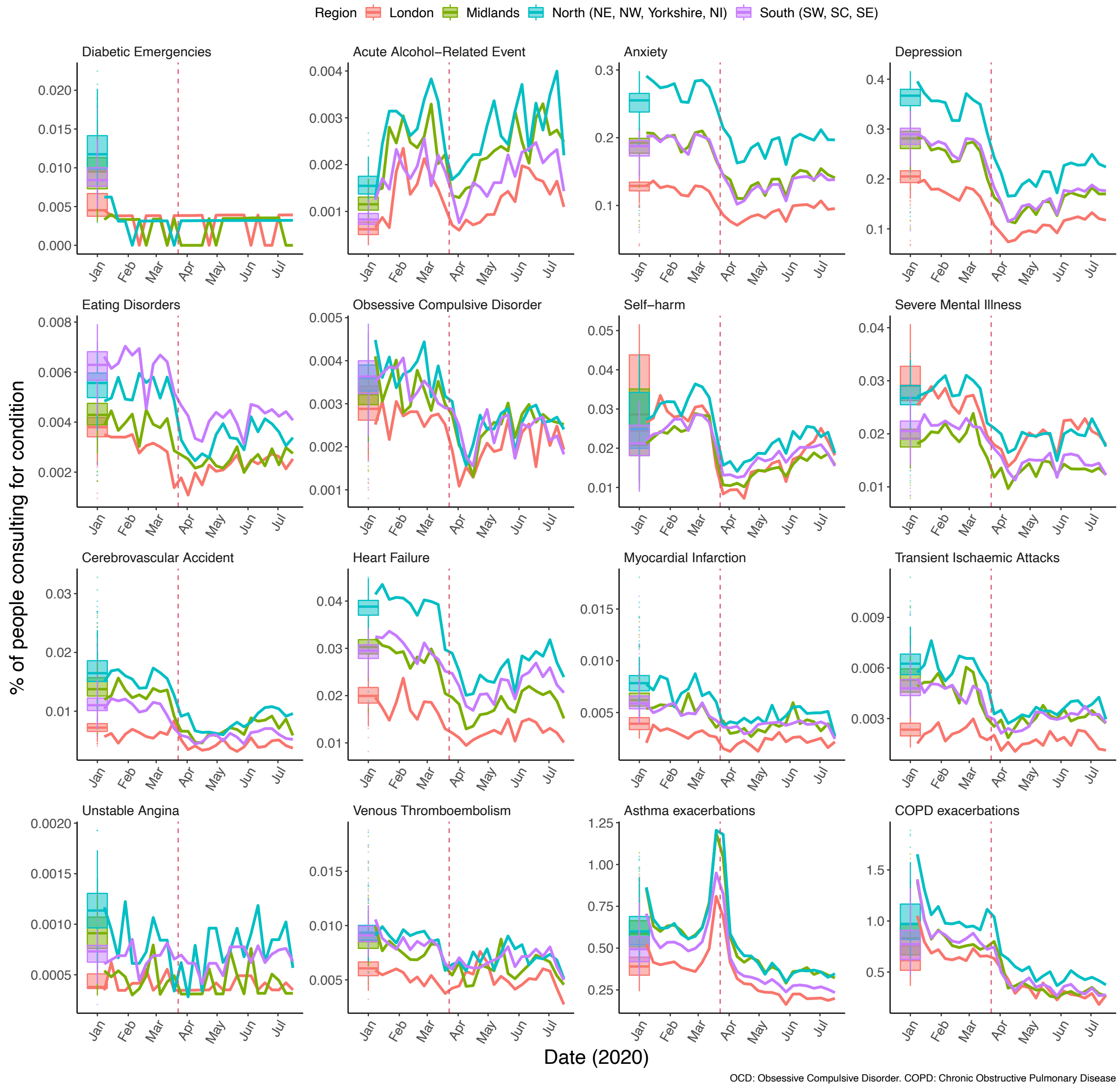

### Text S1

***ITS – the effect of inclusion of different length time series to define the pre-lockdown period***

In our study protocol we planned to use data from 2017 to 2020 to conduct our interrupted time series analysis. We were able to use the full data for all conditions except self-harm, where the data showed a marked and instantaneous level shift in March 2018 (**Figure S5**). Since we hypothesised that this change in recording was likely to be related to a change in primary care coding practice and not reflective of underlying disease burden, these data were excluded from our analysis for self-harm. If we had included this data in the analysis for the definition of pre-restrictions, it would have led to an overestimate of the contact rate for self-harm in March 2020 and therefore overestimated the effect of the restrictions on self-harm consultation. For completeness we present the analysis with data constrained to 2019 onwards for all conditions (**Figure S6**) and the estimates for the effect of the restictions in the immediate reduction in primary care contacts and recovery of contact rates following the introduction of restrictions are shown in a forest plot (**Figure S7**). Pre-restriction was defined as the period up until the first week of March and three weeks of data were excluded, as in the main paper.

***Figure S5 – As Figure 3, full data series plotted. Pre-lockdown period defined as 2017 until 1st March for all conditions (including self-harm; main analysis excluded 2017-2018 data for self-harm).***
Data excluded for 3 weeks between pre- and post-introduction of restriction periods. Odds ratios in B show the relative change in the log odds of contacts for a particular condition on 29th March compared to 1st March 2020.

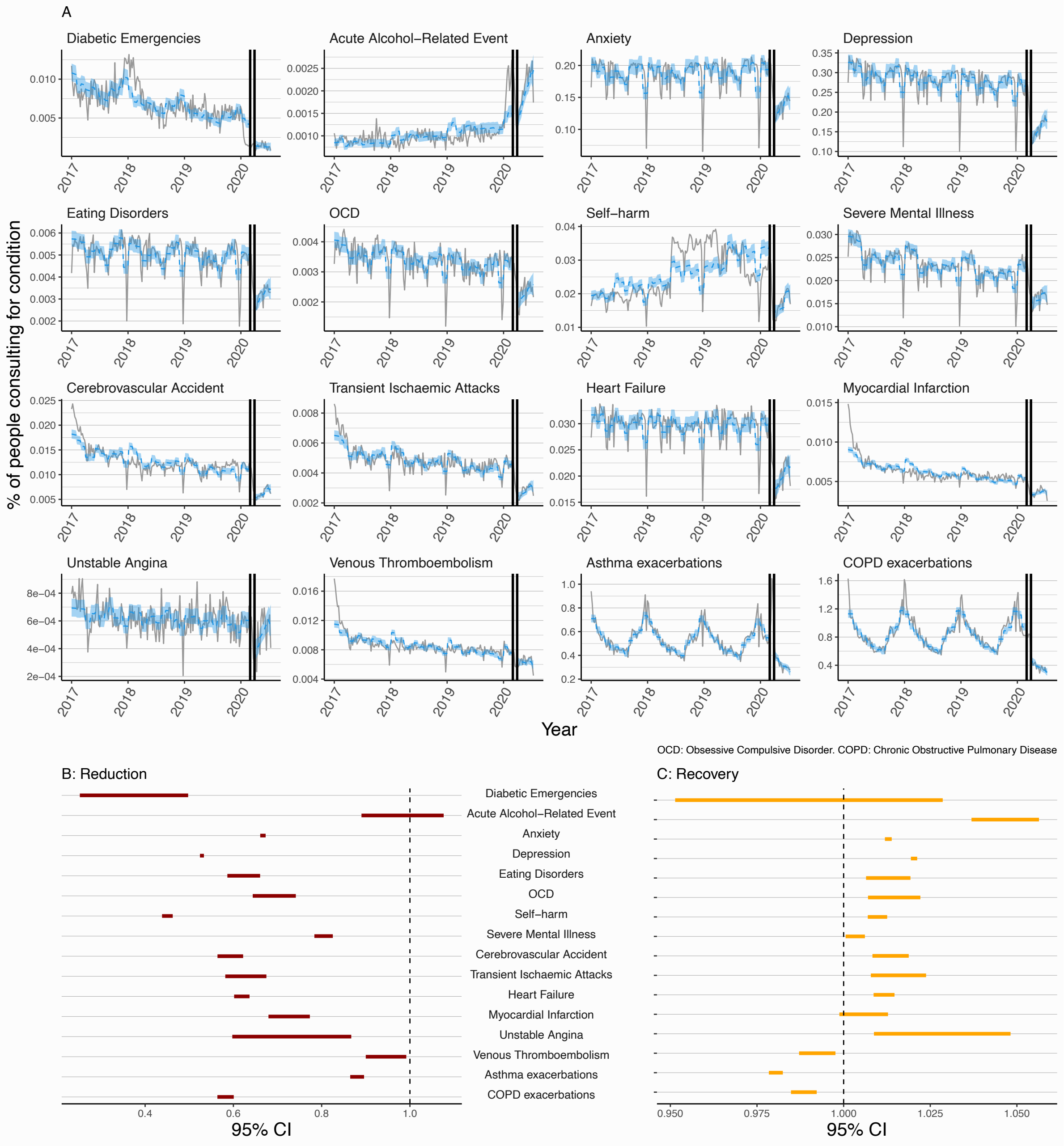

***Figure S6 – As Figure 3, full data series plotted. Pre-lockdown period defined as 2019 until 1st March for all conditions.***

Data excluded for 3 weeks between pre-lockdown and with restrictions periods. Odds ratios in B show the relative change in the log odds of contacts for a particular condition on 29th March compared to 1st March 2020.

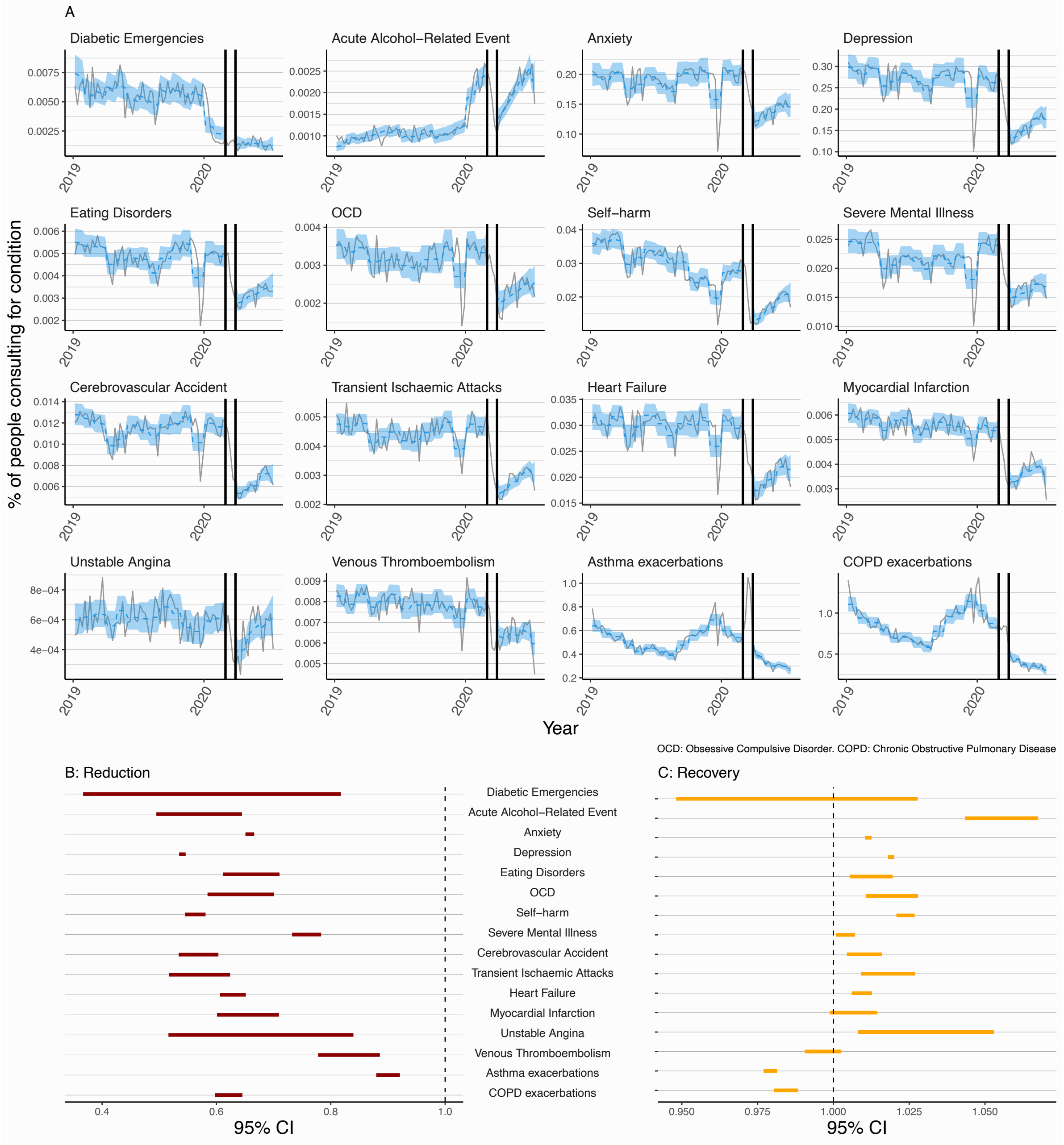

***Figure S7 – A forest plot showing the effect of using different data periods on the estimated effect of the introduction of restrictions on primary care contact behaviour.***

The plot "reduction" shows the odds ratio for the intervention (introduction of restrictions) in our ITS, this shows the relative change in contacts between 29th March and 1st March. "Recovery" shows the effect of time on the odds of contacts in the post-introduciton of restrictions period. Colours indicate analyses using either data from 2017 or data from 2019 as the start of the pre-lockdown period.

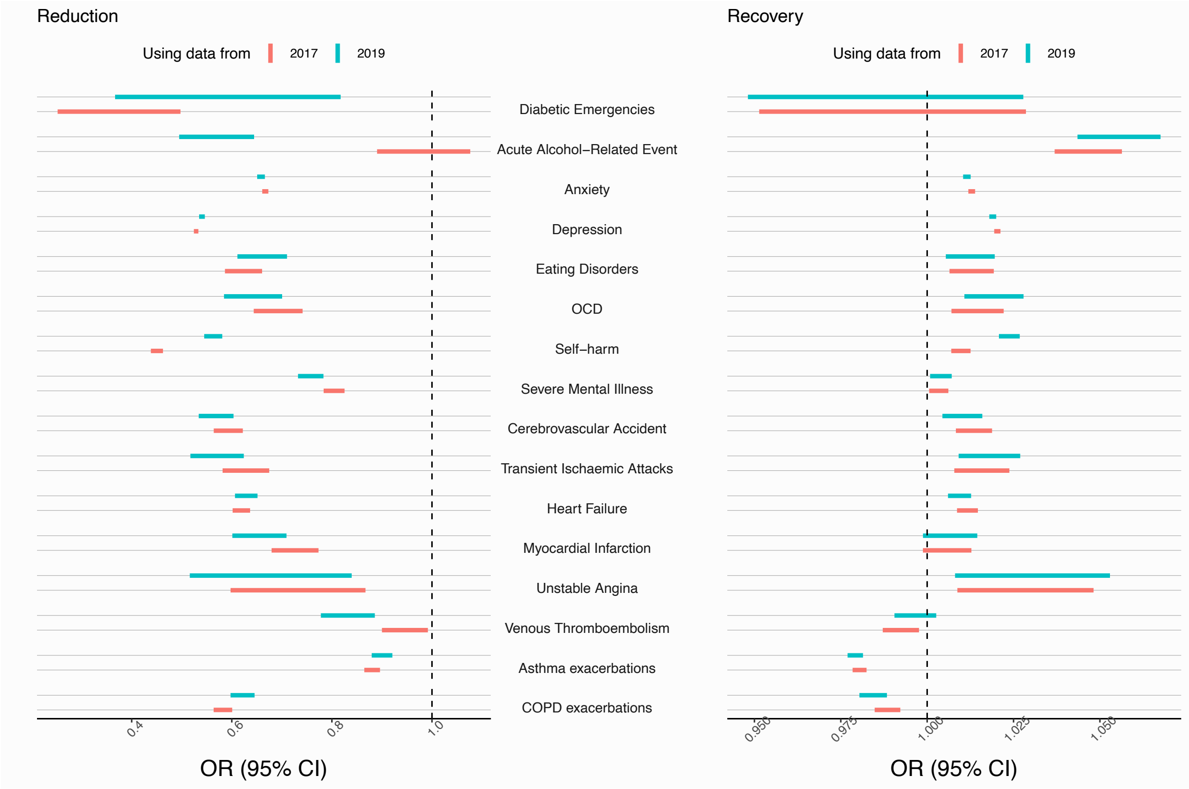

### Text S2

#### *Statistical analysis methods – additional information*

We described all denominator study populations in the first week of January each year (2017-2020) (**Tables 1, S1-S5**). We plotted the percentage of our study populations with contacts for particular conditions in given weeks in 2020 and historical averages for that week (2017-2019). The historical average was the mean of the percentage of the study population consulting for a particular outcome in a given week between 2017 and 2019. We repeated this analysis stratified by age, region, ethnicity, and sex. To protect confidentiality, weekly cell counts were censored as 5 for any value between 0 and 5. If the total number of contacts for a particular condition in a particular strata never exceeded 5 in 2020 then data for these strata were not plotted in our analysis (**Figures S2-S4**).

#### *ITS with variable periods for introduction of restrictions*

We developed an interrupted time series (ITS) model to estimate the effect of restrictions on primary care contacts. The ITS model was a binomial generalised linear model for each condition at time $t$ as defined below:

$$y_t = \beta_0 + \beta_1 T + \beta_2 R_t + \beta_3 T.R_t + \beta_4 Month_t + \beta_5 lagResid_t$$

Where $y_t$ was the proportion of the eligible population with primary care contacts for the condition of interest (e.g. anxiety) in week $t$. The explanatory variables were: 1) time as a linear count of the week of the study ($T$); 2) COVID-19 restrictions, which was a binary variable set to $0$ for the pre-lockdown period and $1$ for the with-restrictions period ($R_t$); and 3) an interaction between these terms ($T.R_t$). The estimate for the coefficient $\beta_2$ is the estimate of the "reduction" in contacts between pre-lockdown and with-restriction periods. The estimate for the coefficient $\beta_3$ is the estimate of the "recovery" in contacts over time with-restrictions, i.e. the effect of a week increase in the with-restrictions period on the log odds of contact for the condition of interest.

The remaining two variables were the calendar month as a categorical variable ($Month_t$) to partially adjust for seasonal variation in primary care contacts. This adjustment was essential for several conditions that show a pronounced seasonal pattern  in contact behaviour in our study (**Figure 2**). The final variable is the lagged residuals from the model ($lagResid_t$). The model was run without this variable and residuals from this reduced model were stored and lagged. These lagged residuals were then used as an explanatory variable in the full model. This was done to adjust for some of the autocorrelation that was present in all our time series, the value of $y_t$ was dependent on the value of $y_{t-1}$.

To display outputs from this model we converted the predicted values (which were predicted log-odds of contact) to a percentage and calculated the 95% confidence interval on the scale of the linear predictor and converted these to a 95% confidence interval for the odds of contacting primary care for a given condition ($\frac{e^x}{1+e^x}$) and multiplying by 100 to convert it to a percentage. We adjusted for overdispersion when calculating these predicted values because we could not assume that at each time-point our $n_i$ Bernoulli trials were indepentally and identically distributed. To do this, we took the Pearson goodness of fit statistic for the full model and divided it by the degrees of freedom in the model. This was used as the dispersion parameter when estimating predicted values instead of the assumed value of 1 from a binomial generalised linear model.

Finally, the structure of the ITS model to estimate the absolute effect of the restrictions on primary care contacts (**Table 3**) was identical except a Poisson model was used and the dynamic population size was included as an offset term. Predicted values from this model were similarly converted from the linear predictor scale (log count) to a count of the number of expected contacts with 95% confidence intervals for a population of 1 million people, which was the exponential of the predicted value for time $t$ divided by the denominator population at time $t$ and multiplied by 1 million.

### Text S3

#### *ITS with variable lockdown periods*

To test the sensitivity of our findings to the choice of pre- and with-restrictions period we repeated the analysis with variable dates for the start of lockdown, and variable lengths of data exclusion to account for adjustment to lockdown in primary care contact behaviour. We varied the last week of pre-lockdown period between either 1st March and 15th March (the week before lockdown announcement in the UK). We varied the period of adjustment-to-restrictions (and therefore excluded data) between 0, 3, 5 or 7 weeks.

*Table S6 – Reduction in contacts: Sensitivity analyses with variable adjustment-to-restrictions periods on primary care contacts comparing periods pre-lockdown and with restrictions*

Results from a sensitivity analysis of variable adjustment-to-restriction periods on the relative change in GP contact behaviour for each health condition. These odds ratios measure the relative change in the log odds of a GP contact with a given health condition between the first week of restrictions period compared to the last week of the pre-lockdown period. All other estimates show strong evidence of a reduction between pre-lockdown and with restrictions periods.

| **Start of behaviour change due to pandemic** | **March 1st** | | | **March 15th** | | | |
|---|---|---|---|---|---|---|---|
| **Duration of adjustment-to-restrictions period excluded from analysis** | **3 weeks (as in Figure 3B, i.e. main analysis)** | **5 weeks** | **7 weeks** | **3 weeks** | **5 weeks** | **7 weeks** | **0 weeks** |
| Diabetic Emergencies | 0.35 (0.25-0.5) | 0.41 (0.29-0.58) | 0.42 (0.29-0.61) | 0.43 (0.3-0.61) | 0.44 (0.3-0.64) | 0.49 (0.33-0.72) | 0.38 (0.27-0.53) |
| Acute Alcohol-Related Events | 0.98 (0.89-1.1) | 1.2 (1.1-1.3) | 1.3 (1.2-1.4) | 1.3 (1.2-1.5) | 1.4 (1.3-1.6) | 1.6 (1.4-1.8) | 1.2 (1.1-1.3) |
| Anxiety | 0.67 (0.66-0.67) | 0.69 (0.68-0.7) | 0.72 (0.71-0.73) | 0.7 (0.69-0.7) | 0.73 (0.72-0.73) | 0.74 (0.73-0.75) | 0.68 (0.68-0.69) |
| Depression | 0.53 (0.52-0.53) | 0.55 (0.55-0.56) | 0.59 (0.59-0.6) | 0.56 (0.55-0.56) | 0.6 (0.59-0.6) | 0.63 (0.62-0.63) | 0.55 (0.54-0.55) |
| Eating Disorders | 0.62 (0.59-0.66) | 0.67 (0.63-0.71) | 0.71 (0.67-0.76) | 0.68 (0.64-0.73) | 0.73 (0.68-0.78) | 0.72 (0.67-0.77) | 0.65 (0.61-0.69) |
| OCD | 0.69 (0.64-0.74) | 0.78 (0.72-0.84) | 0.88 (0.81-0.95) | 0.78 (0.73-0.84) | 0.88 (0.82-0.95) | 0.91 (0.84-0.99) | 0.7 (0.65-0.75) |
| Self-harm | 0.56 (0.54-0.58) | 0.67 (0.65-0.69) | 0.74 (0.71-0.77) | 0.7 (0.68-0.72) | 0.78 (0.75-0.8) | 0.81 (0.78-0.84) | 0.67 (0.65-0.7) |
| Severe Mental Illness | 0.8 (0.78-0.83) | 0.87 (0.85-0.9) | 0.92 (0.9-0.95) | 0.88 (0.86-0.91) | 0.93 (0.9-0.96) | 0.93 (0.9-0.96) | 0.82 (0.8-0.84) |
| Cerebrovascular Accident | 0.59 (0.56-0.62) | 0.62 (0.59-0.65) | 0.66 (0.63-0.7) | 0.64 (0.61-0.68) | 0.69 (0.66-0.73) | 0.74 (0.7-0.79) | 0.65 (0.62-0.68) |
| Transient Ischaemic Attack | 0.63 (0.58-0.67) | 0.7 (0.65-0.76) | 0.77 (0.71-0.84) | 0.73 (0.67-0.78) | 0.81 (0.74-0.88) | 0.86 (0.78-0.93) | 0.67 (0.62-0.72) |
| Heart Failure | 0.62 (0.6-0.64) | 0.65 (0.63-0.67) | 0.69 (0.67-0.71) | 0.66 (0.64-0.68) | 0.7 (0.68-0.72) | 0.75 (0.73-0.78) | 0.64 (0.62-0.66) |
| Myocardial Infarction | 0.72 (0.68-0.77) | 0.78 (0.73-0.84) | 0.86 (0.8-0.92) | 0.82 (0.77-0.88) | 0.9 (0.84-0.97) | 0.99 (0.92-1.1) | 0.79 (0.74-0.84) |
| Unstable Angina | 0.72 (0.6-0.87) | 0.87 (0.72-1) | 0.96 (0.79-1.2) | 0.87 (0.72-1.1) | 0.97 (0.79-1.2) | 1.2 (0.97-1.4) | 0.74 (0.61-0.89) |
| Venous Thromboembolism | 0.94 (0.9-0.99) | 1 (0.96-1.1) | 1 (0.98-1.1) | 1.1 (1-1.1) | 1.1 (1-1.2) | 1.1 (1.1-1.2) | 1 (0.96-1.1) |
| Asthma exacerbations | 0.88 (0.86-0.9) | 0.79 (0.78-0.81) | 0.75 (0.73-0.77) | 0.73 (0.71-0.74) | 0.68 (0.67-0.7) | 0.66 (0.65-0.68) | 0.75 (0.74-0.77) |
| COPD exacerbations | 0.58 (0.56-0.6) | 0.53 (0.51-0.55) | 0.53 (0.51-0.55) | 0.53 (0.51-0.55) | 0.53 (0.51-0.55) | 0.52 (0.5-0.54) | 0.59 (0.57-0.6) |

*Table S7 – Recovery in contacts: Sensitivity analyses with variable adjustment-to-restrictions periods on primary care contacts comparing periods pre-lockdown and with restrictions*

Results from a sensitivity analysis of variable adjustment-to-restrictions periods on the relative effect of time on consultation behaviour for several health conditions with-restrictions. These odds ratios measure the relative effect on the log odds of a GP contact with a given health condition for a unit increase in time (weekly increases) with restrictions.

| **Start of behaviour change due to pandemic** | **March 1st** | | | **March 15th** | | | |
|---|---|---|---|---|---|---|---|
| **Duration of adjustment-to-restrictions period excluded from analysis** | **3 weeks (as in Figure 3C)** | **5 weeks** | **7 weeks** | **3 weeks** | **5 weeks** | **7 weeks** | **0 weeks** |
| Diabetic Emergencies | 0.99 (0.95-1.03) | 0.97 (0.93-1.02) | 0.96 (0.9-1.02) | 0.97 (0.92-1.02) | 0.96 (0.9-1.02) | 0.93 (0.86-1.01) | 0.99 (0.95-1.03) |
| Acute Alcohol-Related Events | 1.05 (1.04-1.06) | 1.03 (1.02-1.04) | 1.03 (1.02-1.04) | 1.02 (1.01-1.03) | 1.01 (1-1.03) | 1 (0.98-1.02) | 1.03 (1.02-1.04) |
| Anxiety | 1.01 (1.01-1.01) | 1.01 (1.01-1.01) | 1.01 (1.01-1.01) | 1.01 (1.01-1.01) | 1.01 (1.01-1.01) | 1.01 (1.01-1.01) | 1.01 (1.01-1.01) |
| Depression | 1.02 (1.02-1.02) | 1.02 (1.02-1.02) | 1.02 (1.02-1.02) | 1.02 (1.02-1.02) | 1.02 (1.01-1.02) | 1.01 (1.01-1.02) | 1.02 (1.02-1.02) |
| Eating Disorders | 1.01 (1.01-1.02) | 1.01 (1-1.02) | 1 (0.99-1.01) | 1.01 (1-1.01) | 1 (0.99-1.01) | 1 (0.99-1.01) | 1.01 (1-1.02) |
| OCD | 1.01 (1.01-1.02) | 1 (1-1.01) | 0.99 (0.98-1) | 1 (1-1.01) | 0.99 (0.98-1) | 0.98 (0.97-1) | 1.01 (1.01-1.02) |
| Self-harm | 1.02 (1.02-1.03) | 1.01 (1.01-1.02) | 1 (1-1.01) | 1.01 (1.01-1.01) | 1 (1-1) | 0.99 (0.99-1) | 1.01 (1.01-1.02) |
| Severe Mental Illness | 1 (1-1.01) | 0.99 (0.99-1) | 0.99 (0.98-0.99) | 0.99 (0.99-1) | 0.99 (0.98-0.99) | 0.98 (0.98-0.99) | 1 (1-1) |
| Cerebrovascular Accident | 1.01 (1.01-1.02) | 1.01 (1.01-1.02) | 1.01 (1-1.02) | 1.01 (1-1.01) | 1 (0.99-1.01) | 0.99 (0.98-1) | 1.01 (1-1.01) |
| Transient Ischaemic Attacks | 1.02 (1.01-1.02) | 1.01 (1-1.02) | 1 (0.98-1.01) | 1 (0.99-1.01) | 0.99 (0.98-1) | 0.98 (0.96-1) | 1.01 (1-1.02) |
| Heart Failure | 1.01 (1.01-1.01) | 1.01 (1.01-1.01) | 1 (1-1.01) | 1.01 (1-1.01) | 1 (1-1.01) | 0.99 (0.98-1) | 1.01 (1.01-1.01) |
| Myocardial Infarction | 1.01 (1-1.01) | 1 (0.99-1.01) | 0.99 (0.98-1) | 0.99 (0.99-1) | 0.98 (0.97-0.99) | 0.96 (0.95-0.98) | 1 (0.99-1.01) |
| Unstable Angina | 1.03 (1.01-1.05) | 1.01 (0.99-1.04) | 1 (0.97-1.03) | 1.01 (0.99-1.04) | 1 (0.97-1.03) | 0.97 (0.93-1.01) | 1.03 (1.01-1.05) |
| Venous Thromboembolism | 0.99 (0.99-1) | 0.98 (0.98-0.99) | 0.98 (0.97-0.98) | 0.98 (0.97-0.99) | 0.97 (0.96-0.98) | 0.96 (0.95-0.97) | 0.99 (0.98-0.99) |
| Asthma exacerbations | 0.98 (0.98-0.98) | 0.99 (0.99-0.99) | 0.99 (0.99-1) | 0.99 (0.99-1) | 1 (1-1) | 1 (1-1.01) | 0.99 (0.99-0.99) |
| COPD | 0.99 (0.98-0.99) | 1 (0.99-1) | 1 (0.99-1) | 1 (0.99-1) | 1 (0.99-1) | 1 (0.99-1.01) | 0.99 (0.98-0.99) |

*Figure S8 – As Figure 3. Pre-lockdown period defined as 2017 until 1st March.*

Data excluded for 3 weeks between pre-lockdown and with-restrictions periods. Odds ratios in B show the relative change in the log odds of contact for a particular condition on 29th March compared to 1st March 2020. Note, this is the same as Figure 3 but is included here for comparison in our sensitivity analysis.

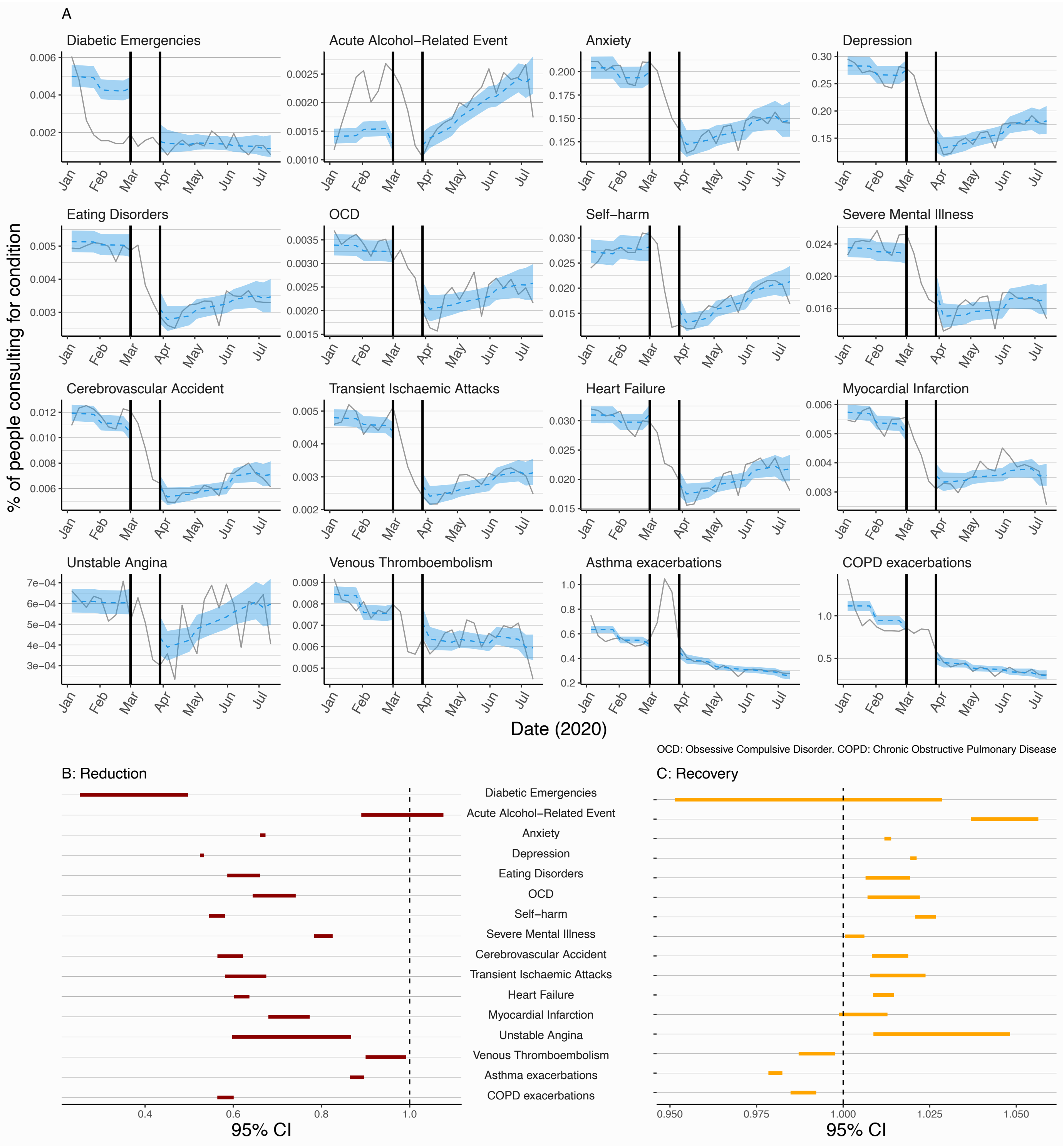

*Figure S9 – As Figure 3. Pre-lockdown period defined as 2017 until 1st March.*

Data excluded for 5 weeks between pre-lockdown and with-restrictions periods. Odds ratios in B show the relative change in the log odds of contact for a particular condition on 12th April compared to 1st March 2020.

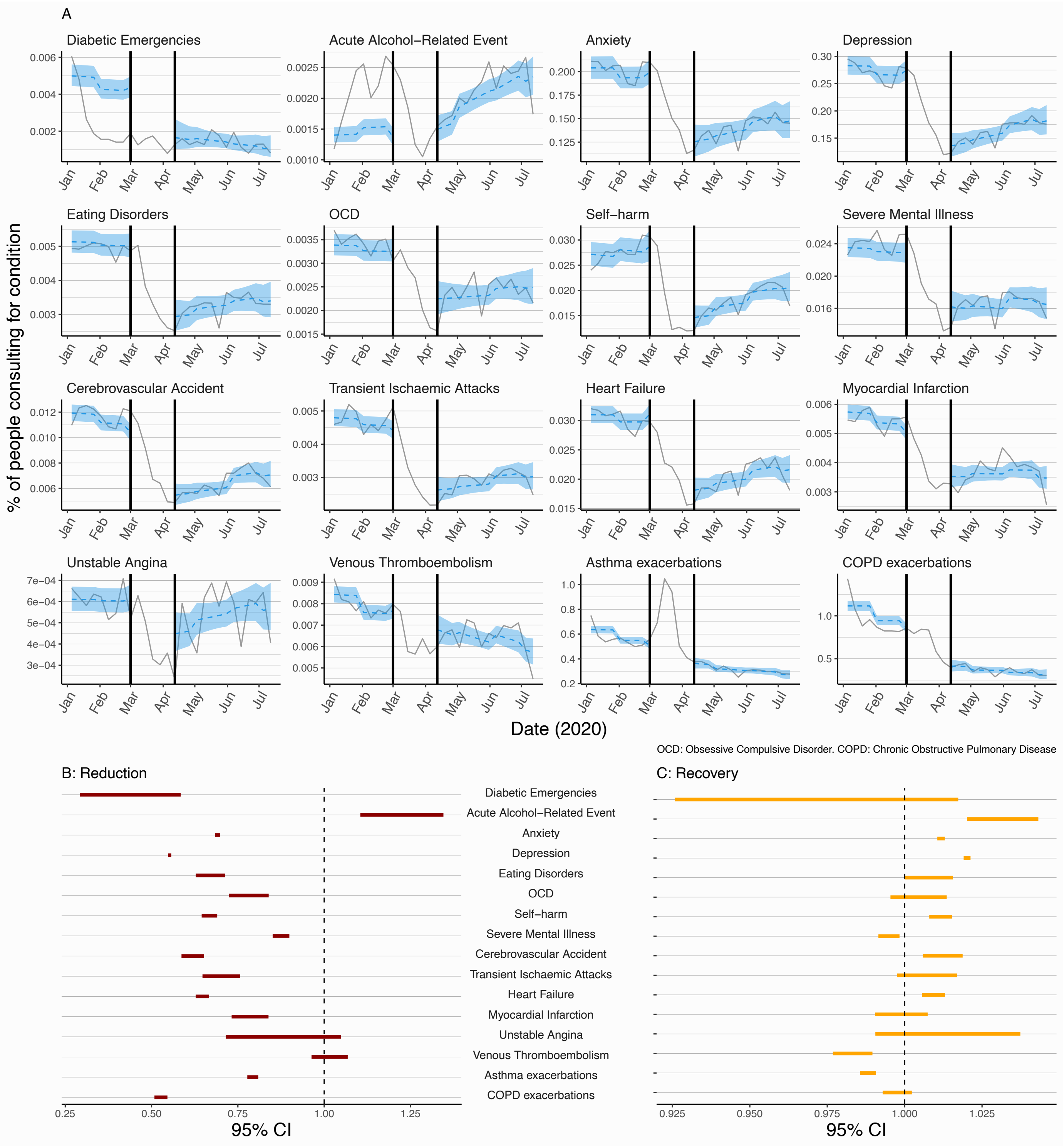

*Figure S10 – As Figure 3. Pre-lockdown period defined as 2017 until 1st March.*

Data excluded for 7 weeks between pre-lockdown and with-restrictions periods. Odds ratios in B show the relative change in the log odds of contact for a particular condition on 26th April compared to 1st March 2020.

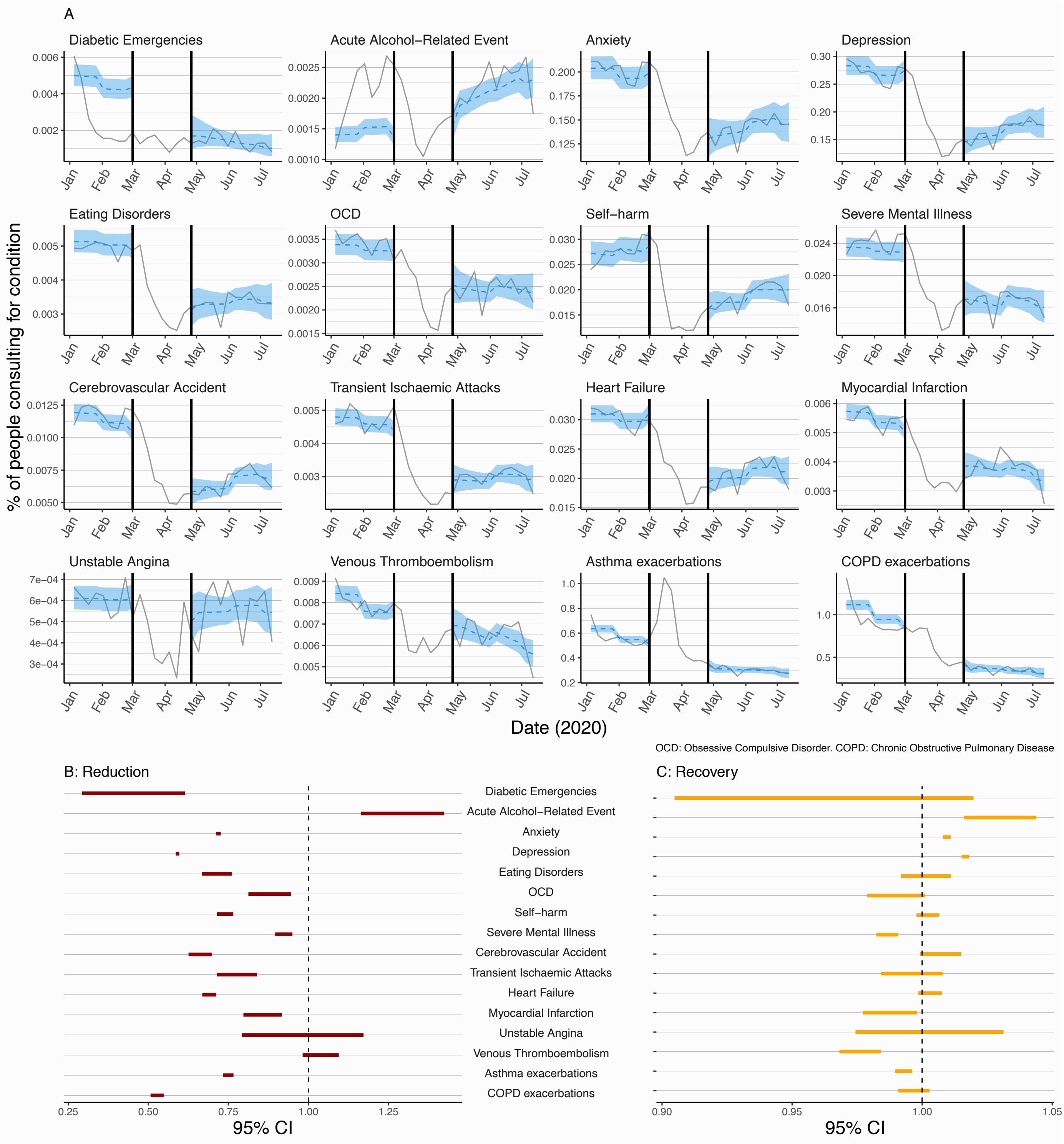

*Figure S11 – As Figure 3. Pre-lockdown period defined as 2017 until 15th March 2020.*
Data excluded for 3 weeks between pre-lockdown and with-restrictions periods. Odds ratios in B show the relative change in the log odds of contact for a particular condition on 12th April compared to 15th March 2020.

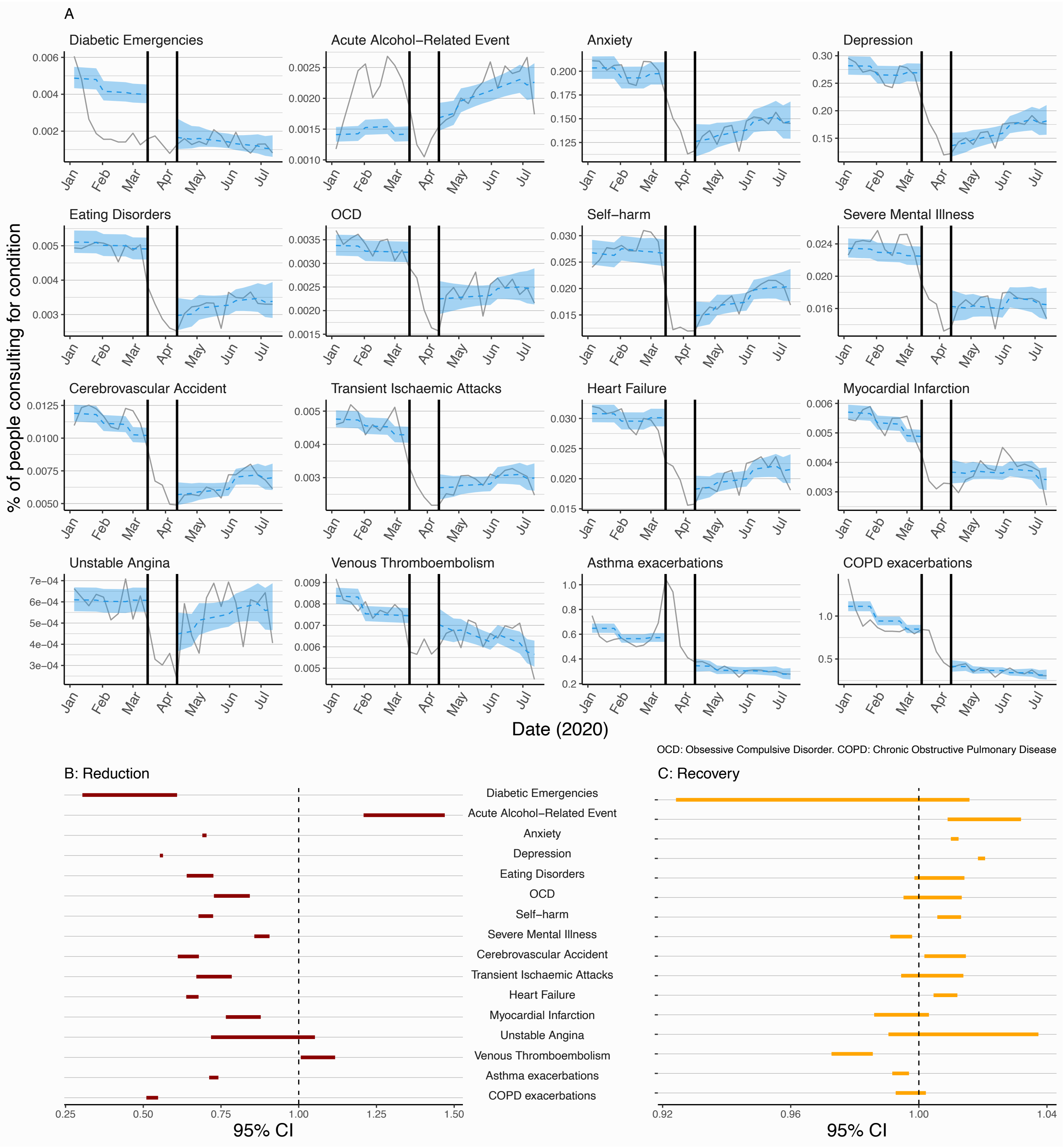

*Figure S12 – As Figure 3. Pre-lockdown period defined as 2017 until 15th March 2020.*

Data excluded for 5 weeks between pre-lockdown and with-restrictions periods. Odds ratios in B show the relative change in the log odds of contact for a particular condition on 26th April compared to 15th March 2020.

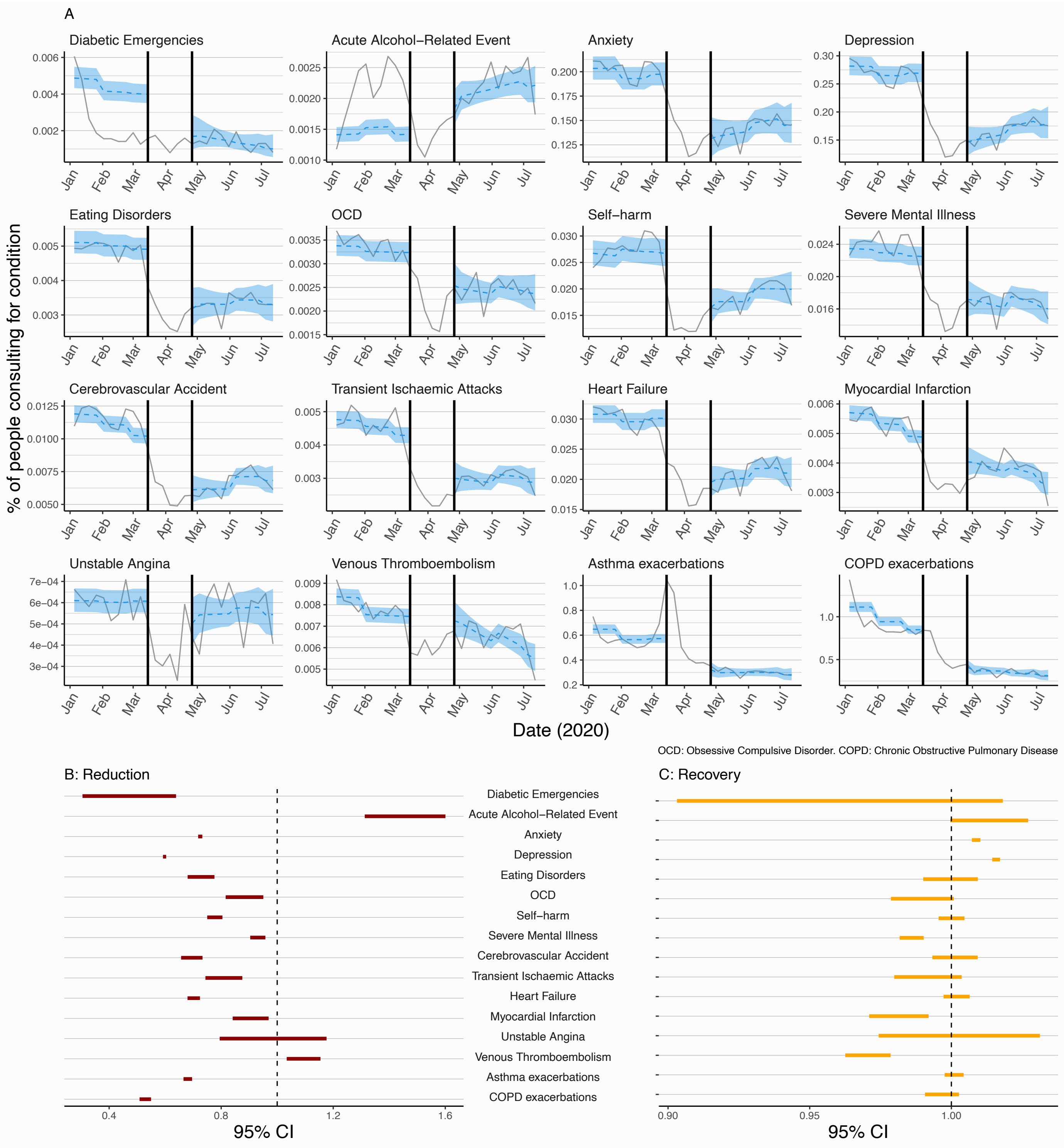

*Figure S13 – As Figure 3. Pre-lockdown period defined as 2017 until 15th March.*

Data excluded for 7 weeks between pre-lockdown and with-restrictions periods. Odds ratios in B show the relative change in the log odds of contact for a particular condition on 10th May compared to 15th March.

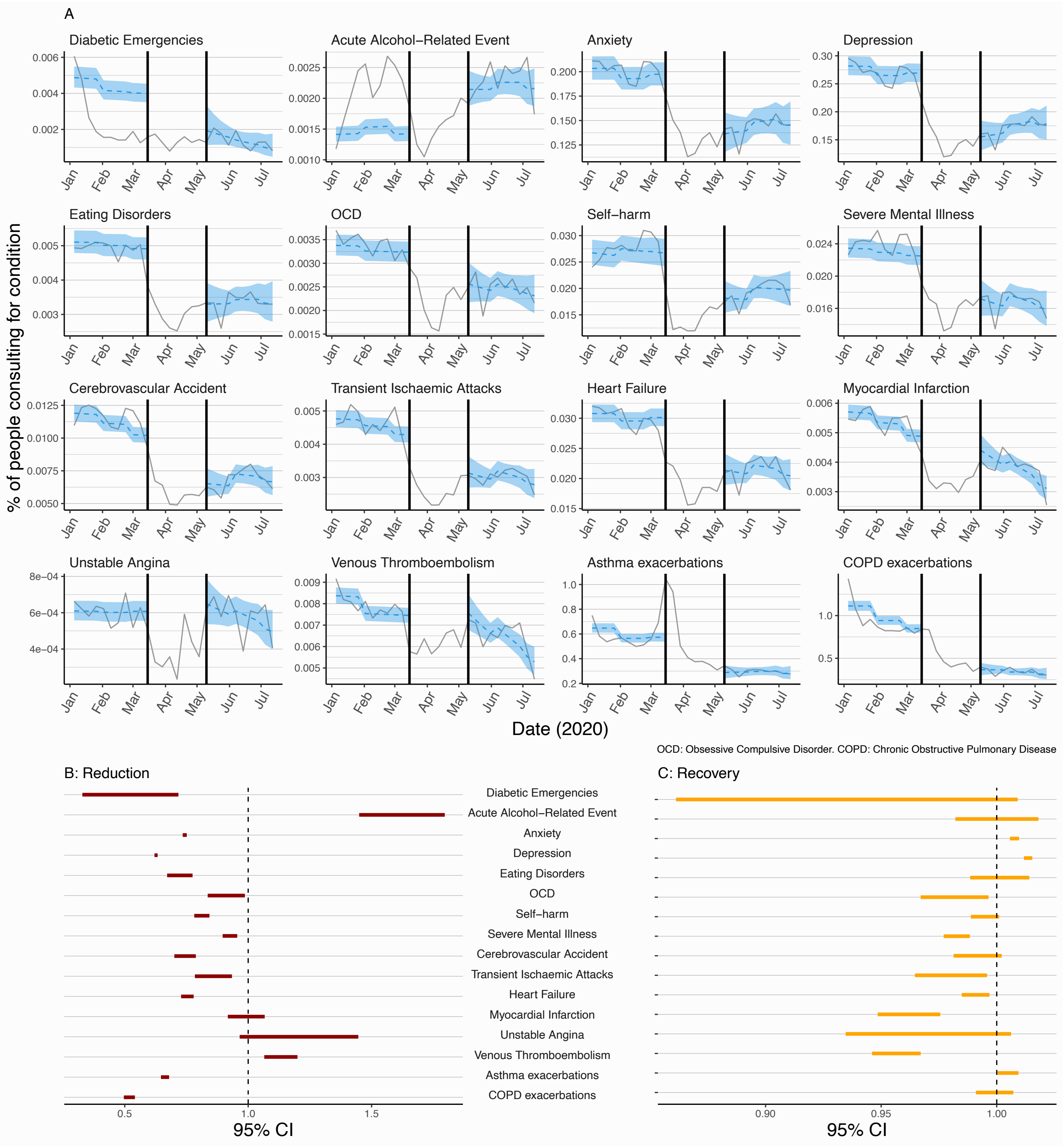

*Figure S14 – As Figure 3. Pre-lockdown period defined as 2017 until 22nd March.*
Data excluded for 0 weeks between pre- and with-restrictions periods. Odds ratios in B show the relative change in the log odds of contact for a particular condition on 29th March compared to 22nd March.

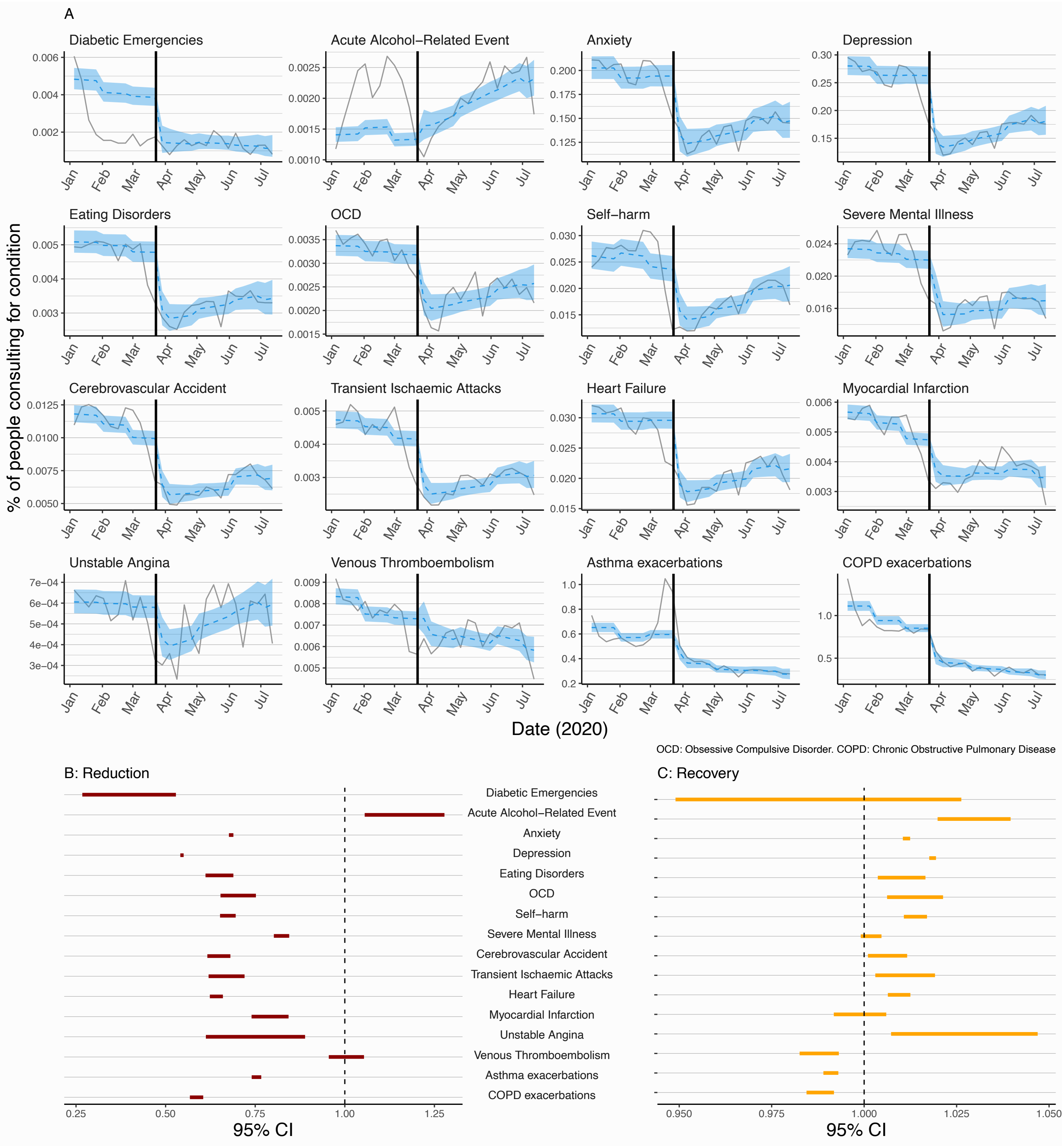

### Text S3

#### *Post-hoc sensitivity analysis: varying diabetic emergencies definition*

Unlike other outcomes, we observed a decline in primary care contacts for diabetic emergencies at the start of 2020, before the implementation of a UK-wide restrictions in March 2020. This may be explained by natural variation, or be artefact due to the small number of conditions. Another explanation may be a delay in recording of hospital records of diabetic emergencies in primary care records (severe diabetic emergencies such as ketoacidoses are likely to lead to hospital admission) due to changes in working patterns in response to the restrictions, leading to inaccurate recording of the dates of contacts and consequently affecting the apparent distribution of contacts.

As a *post-hoc* sensitivity analysis we additionally included records for "non-diabetic hyperglycaemia" in our definition of "diabetic emergencies" (**Table S8**). People with diabetes mellitus were the denominator population for this condition, so it is likely that any hyperglycaemic records (regardless of whether they were labelled 'non-diabetic') were due to diabetes. **Figure S14** shows the results for the two definitions of diabetic emergency (i.e. main and sensitivity analyses) and indicates that even with the inclusion of "non-diabetic hyperglycaemia" as a diabetic emergency (**Figure S14B**) the percentage of diabetic emergency contacts is consistently lower than the historical average (except for spikes in two weeks of June and July 2020). Furthermore, when the definition includes "non-diabetic hyperglycaemia", there was a clear reduction in diabetic emergency contacts across 2020 compared to the 2017-2019 average. We also analysed the trend in all contacts for diabetes in 2020 (routine and emergency codes) (**Figure S14C**), which showed a rapid and sustained decrease beginning shortly before the introduction of UK-wide-restrictions in March 2020 compared to the historical weekly average between 2017-2019 (as is observed across the majority of other conditions).

***Table S8. Main analysis and post-hoc sensitivity analyses with alternative definitions of diabetes condition.***

| Condition | Denominator population | Condition definition |
|---|---|---|
| Diabetes | | |
| Diabetic emergencies (main analysis definition) | All individuals (aged ≥11 years) with prevalent diagnoses of diabetes mellitus at the start of each week of follow-up. Individuals contributed to the study population from the latest of the start of follow-up in the overall population and the date of their first record indicating a diagnosis of diabetes. | Any record of hyperglycaemia, hypoglycaemia, ketoacidosis, or diabetic coma. Multiple records occurring within **seven days** of each other were considered as representing the same event. |
| Diabetic emergenices (*post-hoc* sensitivity analysis) | As above | Any record of hyperglycaemia (recorded as "diabetic" or "non-diabetic"), hypoglycaemia, ketoacidosis, or diabetic coma. Multiple records occurring within **seven days** of each other were considered as representing the same event. |
| All diabetes primary care contacts (*post-hoc* sensitivity analysis) | As above | Any record of a consultation involving diabetes, routine or emergency. Multiple records occurring within **seven days** of each other were considered as representing the same event. |

*Figure S15 – Sensitivity analysis of the definition of diabetic contacts.*
(A) trend of 2020 consultations for diabetic emergencies as defined in the main body of this paper.

(B) a post-hoc sensitivity analysis that classified "non-diabetic hyperglycaemia" as a diabetic emergency since it was recorded in a population of people with diabetes.

(C) a post-hoc sensitivity analysis of all diabetes consultations (emergency and routine) in 2020 compared to a historical average.

Black line, weekly historical average percentage of eligible population consulting (2017-2019, grey lines show the data for 2017, 2018, and 2019). Red line, weekly percentage of eligible percentage consulting in 2020. Red shaded region shows difference with historical average. Red dotted line, introduction of restrictions in UK on March 23rd.

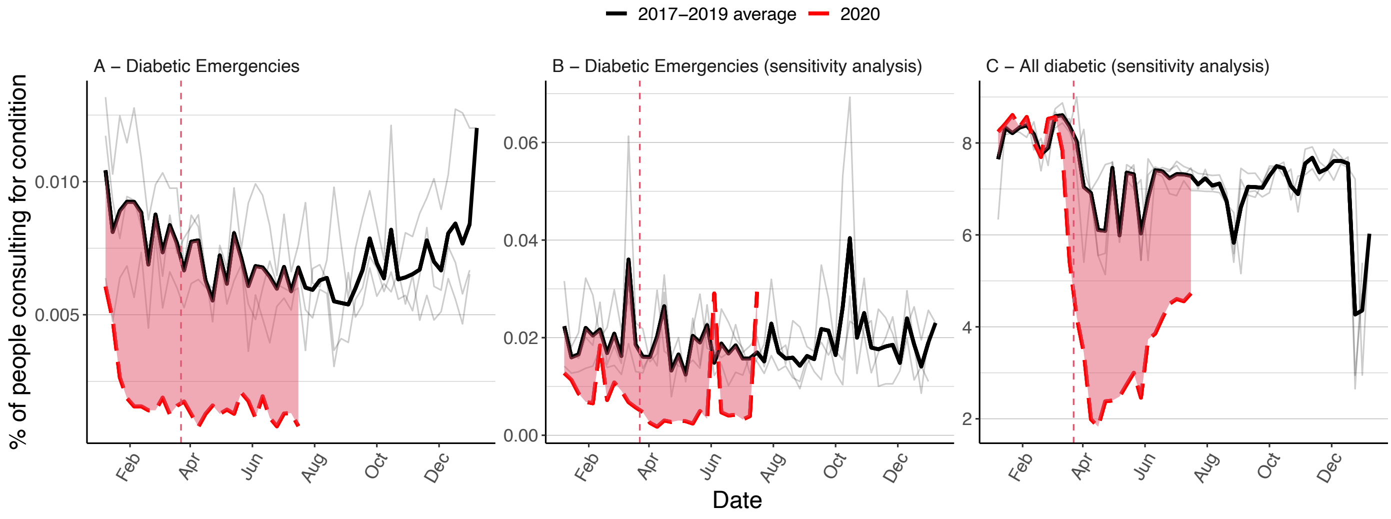
